# From Current-Wave to Longitudinal Risk Prediction: A Leakage-Aware Stacked Ensemble Framework for Adolescent Substance Use Using the ABCD Study

**DOI:** 10.64898/2026.07.08.26357536

**Authors:** Viviana M. Milla-Angeles, Daniel F. Otero-Leon

## Abstract

Adolescent use of alcohol, nicotine, and marijuana remains a major public health concern in the United States. Early identification of youth at elevated risk is critical for prevention before use begins or escalates. We developed and evaluated a longitudinal machine learning framework to predict alcohol, nicotine, and marijuana use at the next observed assessment wave. Data came from the Adolescent Brain Cognitive Development (ABCD) Study Release 6.0. The models incorporated predictors from multiple domains, including demographics, friends, family and community context, mental health, physical health, and prior substance-related behaviors. To reduce information leakage across individuals, we implemented a leakage-aware stacked ensemble. This ensemble combined diverse base learners through out-of-fold predictions and an elastic-net meta-learner. Across all three substances, the lagged stacked ensemble outperformed the cross-sectional stack and all single base learners. Adolescents identified as highest risk showed substantially higher observed rates of substance use than would be expected under random screening. Feature-importance analyses showed that the full longitudinal models were strongly influenced by developmental timing and prior-use history. Analyses restricted to current-wave features revealed distinct substance-specific risk patterns beyond prior-use history and developmental timing. Bootstrap stability analyses identified top-ranked features showing consistent positive predictive relevance across resampled adolescents. These findings suggest that longitudinal, leakage-aware machine learning can generate substance-specific risk estimates to support targeted prevention and screening in adolescent populations.

## 1. Introduction

Substance use during adolescence continues to pose a significant public health challenge in the United States. National surveillance data continue to show substantial use of alcohol, nicotine, and marijuana during adolescence (CDC, 2024; SAMHSA, 2024). These patterns are concerning given that adolescence is a sensitive developmental period during which substance use is associated with adverse cognitive, behavioral, and health outcomes (CDC, 2024; Gray & Squeglia, 2018). Prevention efforts are therefore needed before use becomes established or intensifies.

A central prevention challenge is identifying adolescents who show elevated risk for later alcohol, nicotine, or marijuana use using information available from earlier assessments. Risk is not expressed uniformly across youth, nor is it fully captured by a single assessment wave. Some adolescents have prior observed evidence of use. Others show peer, family, behavioral, emotional, or contextual patterns that may help predict later substance use. Risk identification therefore requires methods that go beyond same-wave correlates and incorporate prior assessments, developmental timing, and lagged risk information.

Most prior studies have treated adolescent substance-use risk as a static process (Trucco, 2020; Green et al., 2024). Much of this work has relied on cross-sectional or short-horizon survey data and conventional statistical approaches to identify risk and protective factors across peer, family, school, and individual domains. These studies have generated valuable evidence about correlates of adolescent substance use. However, they do not capture how risk evolves across time through prior behavior, changing social environments, mental-health variation, and developmental context. In addition, many studies examine substance use as a general outcome, even though risk factors may not operate in the same way across alcohol, nicotine, and marijuana. For example, de Andrés-Sánchez and Belzunegui-Eraso (2022) showed that cannabis use could be explained through combinations of adolescent and contextual factors rather than through any single variable alone, suggesting that substance-specific pathways may be important. These gaps motivate the central research question of this study: Can longitudinal information from prior assessments improve the identification of adolescents at elevated risk for later alcohol, nicotine, and marijuana use, and do the predictive patterns differ across these substances?

To answer this question, the study focuses on three goals: first, to identify adolescents at elevated risk for later observed alcohol, nicotine, and marijuana use; second, to characterize the features and feature combinations associated with elevated risk; and third, to examine whether predictive patterns differ across substances when prior assessments, developmental timing, and lagged risk information are incorporated. To achieve these goals, we develop and evaluate a longitudinal machine-learning framework using repeated observations from the Adolescent Brain Cognitive Development (ABCD) Study, Release 6.0. The proposed framework combines predictors from demographics, friends, family and community context, mental health, physical health, and prior substance-related behaviors. It uses a leakage-aware stacked ensemble architecture that combines diverse base learners through out-of-fold predictions and an elastic-net meta-learner. This reduces information leakage across individuals while allowing risk to be modeled as a developmental process. By modeling alcohol, nicotine, and marijuana separately within a common longitudinal framework, this work moves beyond one-wave risk-factor analyses. It offers a more predictive, interpretable, and developmentally informed approach for adolescent substance-use prevention. The remainder of this paper is organized as follows: Section 2 reviews related work, Section 3 presents the methodology, Section 4 reports the results, and Section 5 discusses the implications, limitations, and future directions.

## 2. Related Work

To clarify the substance-use and methodological foundations of this study, the related work is organized into two sections. The first reviews prior research on adolescent substance-use risk, including traditional risk-factor studies, longitudinal and developmental perspectives, and recent machine-learning applications for substance-use prediction. This section establishes the motivation for a substance-specific and longitudinal approach to risk prediction. The second reviews methodological work on stacked and interpretable healthcare prediction.

### 2.1 Prior Work on Adolescent Substance-Use Risk

Research on adolescent substance use has consistently identified risk and protective factors across individual, family, peer, school, and contextual domains, including impulsivity, sensation-seeking, mental-health symptoms, family instability, reduced parental monitoring, adverse peer environments, and weaker school engagement (Nawi et al., 2021; Öztaş et al., 2018). This literature shows that adolescent substance use is shaped by multiple interacting influences rather than by isolated predictors. Despite this, most studies have relied on cross-sectional designs and conventional statistical methods (de Andrés-Sánchez & Belzunegui-Eraso, 2022; Öztaş et al., 2018; Rodríguez-Ruiz et al., 2021). Although useful for estimating associations, these approaches are less suited to capturing how risk evolves over time.

Longitudinal and developmental research has begun to address this limitation by examining within-individual patterns of stability and change. Rodríguez-Ruiz et al. (2021) noted that few studies have focused directly on developmental change in adolescent substance use, despite the importance of trajectories for understanding vulnerability. Boer et al. (2024) similarly identified adolescence as a critical period for initiation, when heightened risk-taking occurs alongside major neurodevelopmental change, making early identification and prevention especially important. Gray and Squeglia (2018) argued that adolescent substance use should be understood within a developmental framework, given its links to cognitive, behavioral, and neurodevelopmental outcomes. Together, these studies suggest that effective risk identification requires methods capable of capturing developmental change, not only cross-sectional associations.

Machine learning methods in public health have helped move the field beyond linear and single associations. Afzali et al. (2019) compared multiple machine-learning algorithms to predict adolescent alcohol use across cohorts, and Green et al. (2024) used elastic-net modeling in the ABCD Study to identify predictors of early substance-use initiation. More recently, Kim et al. (2025) developed machine-learning models to predict adolescent substance use across three independent national cohorts from South Korea, the United States, and Norway. They used feature-importance and SHAP analyses to identify key contributors to risk. Ünlü et al. (2025) applied multiple feature-selection methods to identify risk factors for several illicit substances in Finland. Their findings showed that machine learning can identify both substance-specific and shared predictors relevant to prevention. These studies support the use of machine learning for adolescent substance-use risk prediction, but relatively few have integrated longitudinal feature construction, leakage-aware modeling, stacked ensemble learning, and substance-specific comparison within a unified framework.

### 2.2 Methodological Precedent for Stacked and Interpretable Healthcare Prediction

Methodologically, this study builds on a broader healthcare risk-prediction literature. Stacked ensembles improve prediction by combining heterogeneous base learners and allowing a meta-learner to integrate complementary predictive signals (Mienye & Sun, 2022). This approach has been applied across several healthcare classification tasks, including diabetes, cardiovascular disease, and liver disease. For diabetes, Reza et al. (2024) showed that stacking classical machine-learning and neural-network approaches improved accuracy and robustness over single models. Acharya et al. (2025) extended this by combining Random Forest, Gradient Boosting, XGBoost, and Support Vector Machines with imbalance handling and SHAP- and LIME-based explainability. For heart disease, Ganie et al. (2025) used stacking and voting ensembles across multiple datasets incorporating SHAP analysis for transparency. For liver disease, Saranya et al. (2025) used a multi-stage stacking framework that combined heterogeneous base classifiers, feature selection, class balancing, and a logistic-regression meta-learner with SHAP-based interpretability. Collectively, these studies support the use of heterogeneous base learners, imbalance-aware modeling, ensemble stacking, and explainable AI for healthcare risk prediction.

Applying this framework to adolescent substance-use risk, however, requires adaptation to the structure of the ABCD longitudinal dataset. Unlike static clinical datasets, the ABCD data include repeated observations from the same adolescents across developmental waves (Hawes et al., 2025). This makes participant-level leakage control, longitudinal feature construction, and prior-use history variables essential. The goal is not only to predict risk, but also to identify the features and feature combinations associated with how that risk develops over time. Therefore, the present study builds on this methodological work by developing a leakage-aware, stacked ensemble pipeline tailored to the ABCD data structure, with alcohol, nicotine, and marijuana modeled as separate outcomes.

## 3. Methodology

This study developed a multi-stage leakage-aware machine-learning pipeline to model adolescent substanceuse risk using longitudinal ABCD 6.0 data. The pipeline was applied separately for alcohol, nicotine, and marijuana and is organized into five phases, illustrated in Figure 1.

**Figure 1.**
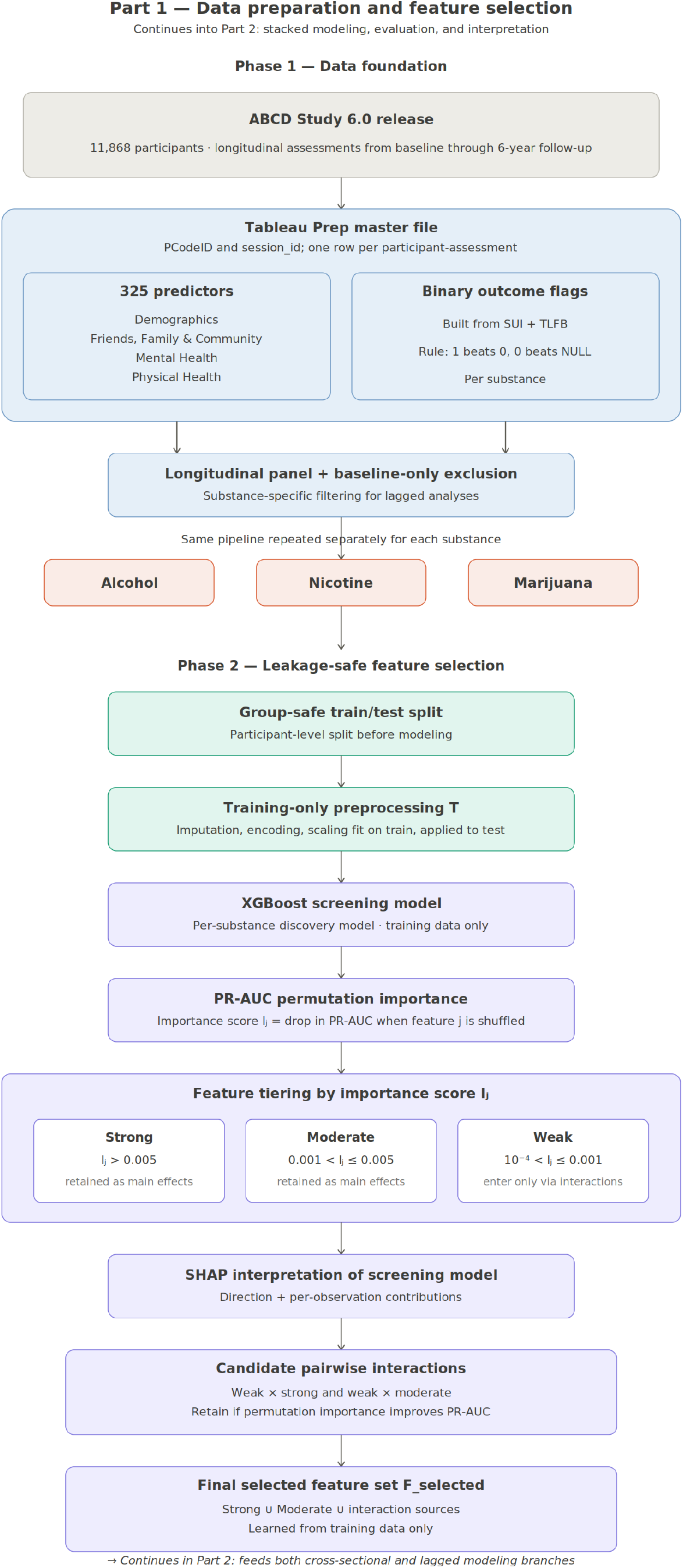
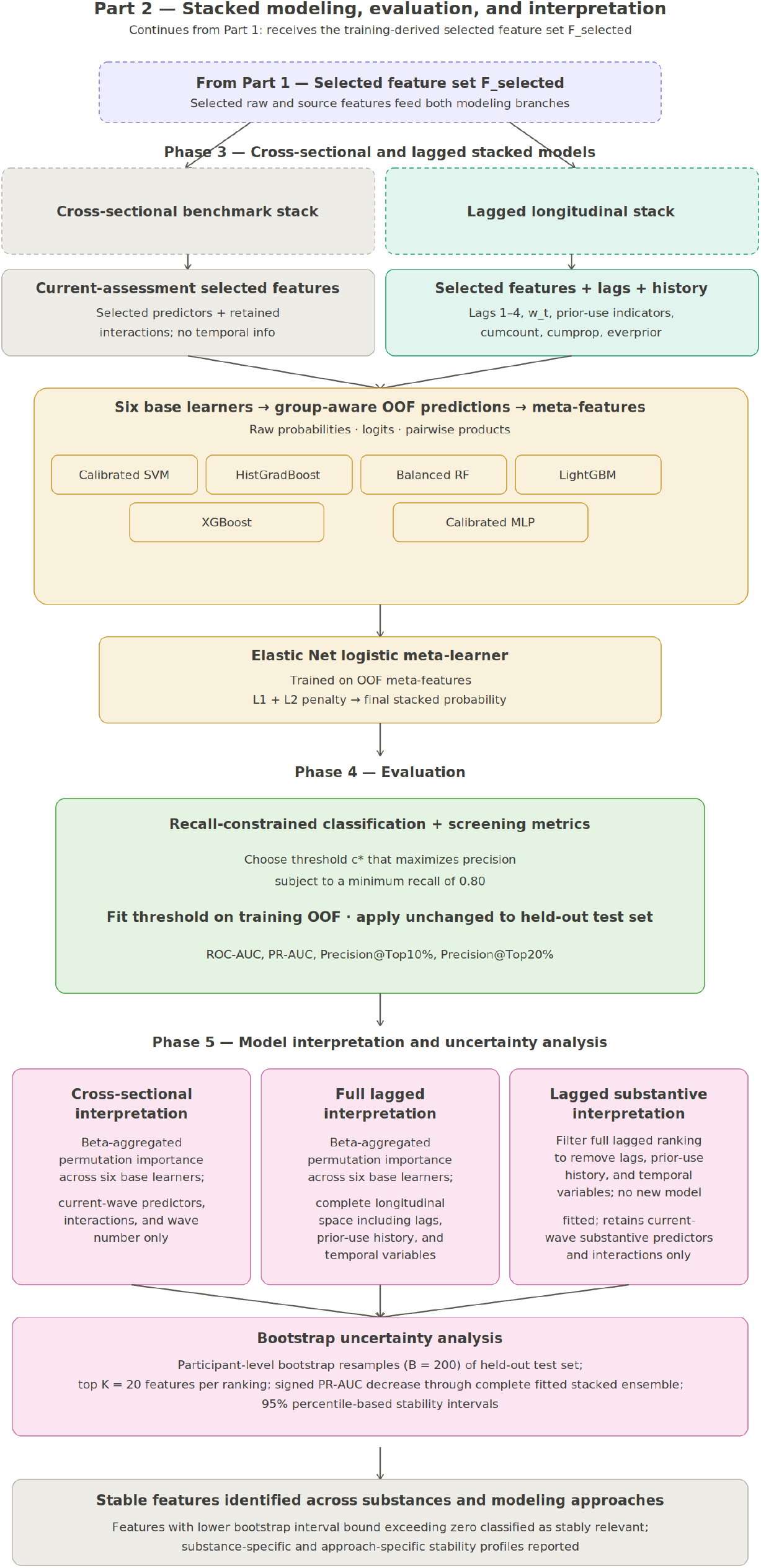
Leakage-aware stacked ensemble pipeline for longitudinal adolescent substance-use risk modeling. The pipeline is organized into five phases across two parts: data integration and leakage-safe feature selection (Phases 1–2), followed by cross-sectional and lagged stacked ensemble modeling with recall-constrained evaluation (Phases 3–4). Model interpretation is performed in three passes (cross-sectional, full lagged, and non-temporal), supplemented by bootstrap uncertainty estimation (Phase 5).

Phase 1 establishes the data foundation by integrating ABCD data and constructing substance-specific binary outcomes. Phase 2 then performs leakage-aware feature selection, applying a group-safe train/test split, training-only preprocessing, and permutation-based importance tiering to identify a clean candidate feature set. Building on this, Phase 3 constructs cross-sectional and lagged stacked ensemble models, combining six heterogeneous base learners with an elastic-net meta-learner. Phase 4 evaluates model performance using recall-constrained threshold calibration applied to held-out test data. Finally, Phase 5 interprets the models through cross-sectional, full lagged, and lagged substantive analyses, supplemented by bootstrap uncertainty estimation.

### 3.1 Phase 1: Data Foundation

Phase 1 establishes the data infrastructure for the entire pipeline. This phase covers the selection and integration of the ABCD 6.0 dataset, the construction of substance-specific outcomes, and the organization of the longitudinal panel structure that all subsequent phases depend on.

#### 3.1.1 Data Integration and Predictor Domains

Tableau Prep was used to assemble a participant-wave longitudinal dataset from multiple ABCD tables (ABCD Study, 2025). The resulting file contained 323 candidate predictors spanning four primary domains: demographics; friends, family, and community; mental health; and physical health. Within the same participant-wave framework, substance-use variables from the Substance Use Interview (SUI) and the Timeline Followback Interview Results (TLFB) were combined. These were used to construct three substance-specific outcome flags for alcohol, nicotine, and marijuana use. Static demographic variables were retained as baseline contextual measures, while repeated measures were preserved longitudinally for downstream modeling. This integrated participant-wave file provided the foundation for all subsequent modeling steps, from feature screening and outcome construction through final model interpretation. A detailed description of the predictor domains and variable distribution is provided in Section 4.1 and Table C1 in the Appendix.

#### 3.1.2 Outcome Definition

Three binary outcomes were defined: Alcohol v. Non Users (at all), Nicotine v. Non Users (at all), and Marijuana (MJ) v. Non Users (at all). The positive class indicated evidence of use of the focal substance; the comparison class consisted of participants with no evidence of use of any substance.

Outcome flags were derived by combining multiple source-level variables from SUI and TLFB. These variables are called raw indicators because they came directly from the source instruments prior to any harmonization, and could differ in format, naming, missingness, and response structure. A participant-wave was coded as positive if any raw indicator showed use, negative if no indicator showed use but at least one explicitly indicated non-use, and missing otherwise. This rule is summarized as “1 beats 0, and 0 beats NULL.”

For participant *i*, wave *t*, and substance *s*, the binary outcome was defined as:

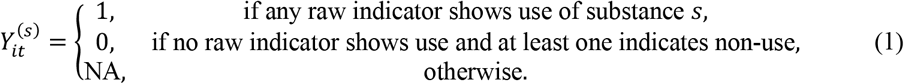

Equation (1) collapses multiple raw substance-use indicators into a single participant-wave label. This outcome serves as the basis for both cross-sectional prediction and prior-history feature construction.

#### 3.1.3 Longitudinal Data Structure and Baseline-Only Exclusion

The analytic dataset was represented as a longitudinal panel:

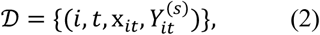

where *i* indexes participants, *t* indexes assessment waves, x_*it*_ is the predictor vector for participant *i* at wave *t*, and 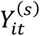 is the binary outcome for substance *s*.

The feature vector is:

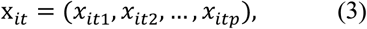

where *p* = 323 is the number of candidate predictors.

Because the primary models required prior-wave information, participants with only baseline data were excluded from the lagged modeling dataset. This exclusion removed 334 cases for alcohol, 320 for nicotine, and 311 for marijuana.

#### 3.1.4 Phase 1: Output and Transition

Together, Sections 3.1.1 through 3.1.3 produced three substance-specific longitudinal datasets for alcohol, nicotine, and marijuana. Each dataset was structured as a participant-wave panel with harmonized predictors and a substance-specific binary outcome. These datasets formed the input to Phase 2.

Before feature selection, a key methodological risk had to be addressed. To prevent leakage, a participant-level group-safe split was applied before any feature-selection decisions were made. All preprocessing, screening, and feature-selection steps were fit on training data only and applied to the held-out test data.

**Table 1.** Notation for the Cross-Sectional and Longitudinal Stacked Ensemble Modeling Framework. Table 1 defines the mathematical notation used throughout the methodology, with symbols grouped by methodological component for ease of reference.

1. Indexing and data structure
| Symbol | Definition |
| --- | --- |
| $i$ | Participant index. |
| $t$ | Wave or visit index. |
| $\tau$ | Prior-wave index used in cumulative summaries over assessments occurring before wave $t$. |
| $r \equiv (i, t)$ | Observation index; observation $r$ corresponds to participant $i$ at wave $t$. |
| $s$ | Substance-specific outcome index, where $s \in \{\text{alcohol}, \text{nicotine}, \text{marijuana}\}$. |
| $\eta$ | Model-design index, where $\eta \in \{\text{CS}, \text{LAG}\}$ denotes the cross-sectional or lagged stacked ensemble. |
| $j$ | Predictor or feature index. |
| $a, b$ | Predictors used to construct an interaction term, or base-learner outputs used to construct a pairwise meta-feature product. |
| $u$ | Lag-order index for predictor features, where $u = 1, \dots, 4$. |
| $k$ | Prior-outcome lag index, where $k = 1, \dots, 4$. |
| $q$ | Cross-validation fold index. |
| $v$ | Meta-feature index in the Elastic Net meta-learner coefficient vector. |
| $N$ | Number of participants in the analytic dataset. |
| $n$ | Number of participant-wave observations used in a model or importance calculation. |
| $p$ | Number of candidate predictors before feature screening. |
| $p_{\text{selected}}^{(s, \eta)}$ | Number of selected current-wave predictors retained for substance $s$ and model design $\eta$. |
| $p_{\eta}^{(s)}$ | Total number of predictors included in the final feature space for substance $s$ under model design $\eta$. |
| $d$ | Number of meta-features supplied to the stacking meta-learner. |
| $D$ | Complete longitudinal analytic dataset. |
| $D_{\text{train}}$ | Training dataset after the group-safe participant split. |
| $D_{\text{test}}$ | Held-out test dataset after the group-safe participant split. |
| $D_{\text{lagged}}^{(s)}$ | Lagged dataset for substance $s$, containing observations with at least one observed prior assessment. |
| $\mathcal{I}$ | Complete set of participant indices. |
| $\mathcal{I}_{\text{train}}$ | Set of participants assigned to the training partition. |
| $\mathcal{I}_{\text{test}}$ | Set of participants assigned to the held-out test partition. |
| $G_i$ | Participant-group identifier for participant $i$, corresponding to PCodeID. |

2. Predictors, outcomes, and preprocessing
| Symbol | Definition |
| --- | --- |
| $\mathbf{x}_{it}$ | Vector of selected current-wave predictors for participant $i$ at wave $t$. |
| $x_{itj}$ | Value of predictor $j$ for participant $i$ at wave $t$. |
| $\mathbf{x}_{i,t-u}$ | Vector of predictors for participant $i$ at predictor lag $u$, corresponding to a prior observed assessment. |
| $X_{i,t-u}^{(j)}$ | Lagged value of predictor $j$ for participant $i$ at predictor lag $u$. |
| $\mathbf{x}_{it}^{\text{lag}}$ | Complete lagged feature vector for participant $i$ at wave $t$, including current-wave predictors, lagged predictors, prior-use history variables, temporal variables, and reconstructed interactions. |
| $\mathbf{x}_r$ | Predictor vector for observation $r$, where $r \equiv (i, t)$. |
| $\mathbf{x}_r^{\text{lag}}$ | Complete lagged predictor vector for observation $r$. |
| $Y_{it}^{(s)}$ | Binary outcome for substance $s$ for participant $i$ at wave $t$. |
| $Y_r^{(s)}$ | Binary outcome for observation $r$ and substance $s$. |
| $y_r^{(s)}$ | Observed binary outcome value for observation $r$ and substance $s$. |
| $P(Y_{it}^{(s)} = 1 \mid \mathbf{x}_{it})$ | Conditional probability of the substance-specific outcome for participant $i$ at wave $t$, given current-wave predictors. |
| $X_{\text{train}}$ | Training feature matrix before preprocessing. |
| $X_{\text{test}}$ | Test feature matrix before preprocessing. |
| $\mathbf{y}_{\text{train}}^{(s)}$ | Training outcome vector for substance $s$. |
| $\mathbf{y}_{\text{test}}^{(s)}$ | Test outcome vector for substance $s$. |
| $\mathcal{T}$ | Preprocessing transformation fitted using the training data only. |
| $X_{\text{train}}^{\text{prep}}$ | Preprocessed training feature matrix. |
| $X_{\text{test}}^{\text{prep}}$ | Preprocessed test feature matrix. |

3. Feature screening and interaction selection
| Symbol | Definition |
| --- | --- |
| $\hat{f}_{\text{screen}}^{(s)}$ | XGBoost feature-screening model trained for substance $s$. |
| $\hat{p}_r^{\text{screen}(s)}$ | Predicted probability from the XGBoost screening model for observation $r$ and substance $s$. |
| $I_j^{\text{screen}(s)}$ | Screening-stage permutation-importance score for predictor $j$ and substance $s$. |
| $I_{(1)}^{\text{screen}(s)}, I_{(2)}^{\text{screen}(s)}, \dots, I_{(p)}^{\text{screen}(s)}$ | Screening-stage permutation-importance scores for substance $s$, ordered from largest to smallest. |
| $I_{ab}$ | Permutation-importance score for the interaction between predictors $a$ and $b$ during interaction screening for substance $s$. |
| $F_{\text{strong}}^{(s,\eta)}$ | Set of strong features retained from screening for substance $s$ and model design $\eta$. |
| $F_{\text{moderate}}^{(s,\eta)}$ | Set of moderate features retained from screening for substance $s$ and model design $\eta$. |
| $F_{\text{weak}}^{(s,\eta)}$ | Set of weak features identified during screening for substance $s$ and model design $\eta$. |
| $F_{\text{selected}}^{(s,\eta)}$ | Final selected current-wave feature set retained for predictive modeling for substance $s$ and model design $\eta$. |
| $F_{\text{interaction sources}}^{(s,\eta)}$ | Source variables required to reconstruct retained interaction terms for substance $s$ and model design $\eta$. |
| $F_{\text{interactions}}^{(s,\eta)}$ | Set of reconstructed interaction terms included in the fitted model for substance $s$ and model design $\eta$. |
| $X_{it,a}$ | Encoded or transformed value of predictor $a$ for participant $i$ at wave $t$. |
| $Z_{it}^{(a,b)}$ | Reconstructed interaction term between predictors $a$ and $b$ for participant $i$ at wave $t$. |
| $X_{\text{train}}^{\text{selected}(s,\eta)}$ | Training matrix restricted to the selected feature space for substance $s$ and model design $\eta$. |
| $X_{\text{test}}^{\text{selected}(s,\eta)}$ | Test matrix restricted to the selected feature space for substance $s$ and model design $\eta$. |

4. Stacked ensemble and meta-learning
| Symbol | Definition |
| --- | --- |
| $\mathcal{M}$ | Set of base learners included in the stacking ensemble. |
| $M$ | Number of base learners in the stacking ensemble; $M = 6$. |
| $\hat{f}_m^{(s,\eta)}$ | Fitted base learner $m$ for substance $s$ and model design $\eta$. |
| $\hat{f}_m^{-q,(s,\eta)}$ | Base learner $m$ fitted excluding cross-validation fold $q$, used to generate out-of-fold predictions. |
| $\hat{p}_{r,m}^{(s,\eta)}$ | Predicted probability for observation $r$ from fitted base learner $m$. |
| $\hat{p}_{r,m}^{\text{OOF}(s,\eta)}$ | Out-of-fold predicted probability for observation $r$ from base learner $m$. |
| $\mathbf{p}_r^{\text{OOF}(s,\eta)}$ | Vector of out-of-fold base-learner probabilities for observation $r$. |
| $\ell_{r,m}^{(s,\eta)}$ | Logit-transformed base-learner probability for observation $r$ from base learner $m$. |
| $\epsilon$ | Small positive constant used to prevent undefined or infinite logit values when predicted probabilities approach 0 or 1. |
| $Q_{r,(a,b)}^{(s,\eta)}$ | Pairwise product of selected base-learner prediction outputs $a$ and $b$ for observation $r$. |
| $\mathbf{z}_r^{(s,\eta)}$ | Meta-feature vector for observation $r$, including base probabilities, logit transformations, pairwise products, and summary statistics. |
| $\hat{g}^{(s,\eta)}$ | Elastic Net Logistic Regression meta-learner for substance $s$ and model design $\eta$. |
| $\beta_0$ | Meta-learner intercept. |
| $\boldsymbol{\beta}$ | Meta-learner coefficient vector. |
| $\beta_v$ | Elastic Net coefficient for meta-feature $v$. |
| $\lambda$ | Elastic Net regularization strength. |
| $\alpha$ | Elastic Net mixing parameter. |
| $\sigma(\cdot)$ | Logistic sigmoid function. |
| $L(\beta_0, \boldsymbol{\beta})$ | Elastic Net Logistic Regression objective function. |
| $\ \boldsymbol{\beta}\ _1$ | $L_1$ norm of the meta-learner coefficient vector, used for sparsity in Elastic Net regularization. |
| $\ \boldsymbol{\beta}\ _2^2$ | Squared $L_2$ norm of the meta-learner coefficient vector, used for shrinkage in Elastic Net regularization. |
| $\hat{p}_r^{(s,\eta)}$ | Final predicted probability for observation $r$ from the fitted stacked ensemble. |
| $\hat{Y}_r^{(s,\eta)}(c)$ | Predicted class for observation $r$ after applying classification threshold $c$. |

5. Beta-aggregated permutation importance
| Symbol | Definition |
| --- | --- |
| $R_\pi$ | Number of permutation repetitions used to estimate base-learner feature importance; $R_\pi = 5$. |
| $\nu$ | Permutation-repeat index, where $\nu = 1, \dots, R_\pi$. |
| $\pi_j$ | Permutation operation applied to feature $j$, leaving all other predictors unchanged. |
| $\text{AP}_m^{(s,\eta)}$ | Baseline test-set PR-AUC for base learner $m$, substance $s$, and model design $\eta$. |
| $\text{AP}_{m,j,\nu}^{\pi(s,\eta)}$ | Test-set PR-AUC for base learner $m$ after permuting feature $j$ in permutation repeat $\nu$. |
| $J_{mj}^{(s,\eta)}$ | Absolute mean PR-AUC decrease after permutation of feature $j$ in base learner $m$. |
| $\tilde{J}_{mj}^{(s,\eta)}$ | Within-learner normalized permutation importance of feature $j$ for base learner $m$. |
| $\kappa_m^{(s,\eta)}$ | Absolute applicable Elastic Net coefficient representing the contribution of base learner $m$, obtained from the raw-probability coefficient block or, when all raw-probability coefficients are zero, from the logit-output coefficient block. |
| $\omega_m^{(s,\eta)}$ | Normalized coefficient-based weight assigned to base learner $m$ during beta aggregation. |
| $J_{j,\beta}^{(s,\eta)}$ | Beta-aggregated permutation-importance score for feature $j$. |
| $P_{j,\beta}^{(s,\eta)}$ | Percentage contribution of feature $j$ to total beta-aggregated importance within substance $s$ and model design $\eta$. |
| $\hat{\mathbf{p}}_m^{(s,\eta)}$ | Predicted-probability vector produced by fitted base learner $m$ on the held-out test set for substance $s$ and model design $\eta$. |
| $\hat{\mathbf{p}}_{mj,\nu}^{\pi(s,\eta)}$ | Predicted-probability vector produced by fitted base learner $m$ after permuting feature $j$ in permutation repeat $\nu$, for substance $s$ and model design $\eta$. |

6. Lagged and prior-history features
| Symbol | Definition |
| --- | --- |
| $w_t$ | Numeric wave-position variable, represented in the dataset as wave_num. |
| $\text{prev}_{it}^{(s,k)}$ | Prior-use indicator for substance $s$, corresponding to observed outcome status at prior-outcome lag $k$ before wave $t$. |
| $\text{cumcount}_{it}^{(s)}$ | Cumulative count of prior observed substance-use occurrences for substance $s$. |
| $\text{priorwavecount}_{it}$ | Number of observed prior waves available for participant $i$ at wave $t$. |
| $\text{cumprop}_{it}^{(s)}$ | Cumulative proportion of prior observed waves with substance use for substance $s$. |
| $\text{everprior}_{it}^{(s)}$ | Indicator of any prior observed substance use for substance $s$. |
| $\text{history}_{it}^{(s)}$ | Vector of prior-use history variables for substance $s$, including prior-use indicators, cumulative count, cumulative proportion, and ever-prior-use indicator. |
| $F_{\text{lagged}}^{(s)}$ | Complete feature set used in the fitted lagged model for substance $s$, including current-wave predictors, reconstructed interactions, lagged predictors, prior-use history variables, and temporal variables. |
| $F_{\text{lags}}^{(s)}$ | Set of lagged predictor variables in the fitted lagged model for substance $s$, including predictor values from Lag1 through Lag4. |
| $F_{\text{history}}^{(s)}$ | Set of substance-specific prior-use history variables, including prev1–prev4, cumcount_prior, cumprop_prior, and ever_prior. |
| $F_{\text{temporal}}$ | Set of temporal variables used in the lagged model: wave_num and prior_wave_count. |
| $X_{\text{train}}^{\text{lagged}(s)}$ | Complete lagged training matrix for substance $s$. |
| $X_{\text{test}}^{\text{lagged}(s)}$ | Complete lagged test matrix for substance $s$. |

7. Thresholding and evaluation
| Symbol | Definition |
| --- | --- |
| $\mathbb{I}(\cdot)$ | Indicator function equal to 1 when the stated condition is true and 0 otherwise. |
| $c$ | Candidate classification threshold. |
| $c^*$ | Recall-constrained classification threshold selected using training out-of-fold predictions. |
| $TP(c)$ | Number of true positives at threshold $c$. |
| $FP(c)$ | Number of false positives at threshold $c$. |
| $FN(c)$ | Number of false negatives at threshold $c$. |
| $S_q$ | Set of test observations in the top $q\%$ of predicted risk. |
| $\vert S_q\vert $ | Number of observations in the top $q\%$ predicted-risk set. |

8. Non-temporal, non-lagged substantive feature analysis
| Symbol | Definition |
| --- | --- |
| $F_{\text{exclude}}^{(s)}$ | Set of lagged, prior-history, and temporal variables removed from the full lagged beta-aggregated ranking for substance $s$. |
| $F_{\text{SUB}}^{(s)}$ | Retained substantive feature subset obtained by filtering $F_{\text{lagged}}^{(s)}$ to remove $F_{\text{exclude}}^{(s)}$; includes current-wave substantive predictors and retained current-wave interaction terms. |
| $P_{j,\text{SUB}}^{(s)}$ | Percentage contribution of retained substantive feature $j$, recalculated within $F_{\text{SUB}}^{(s)}$. |

9. Bootstrap stability intervals for stacked-model permutation importance
| Symbol | Definition |
| --- | --- |
| $K$ | Number of top-ranked features selected from a beta-aggregated ranking for stacked-model bootstrap stability evaluation; $K = 20$. |
| $B$ | Number of participant-level bootstrap replicates; $B = 200$. |
| $\zeta$ | Bootstrap replicate index, where $\zeta = 1, \dots, B$. |
| $\mathcal{I}_{\text{test}}^{*(\zeta)}$ | Bootstrap sample of test-set participant indices in replicate $\zeta$, sampled with replacement. |
| $D_{\text{test}}^{*(\zeta)}$ | Bootstrap test dataset formed by retaining all participant-wave observations belonging to participants sampled in $\mathcal{I}_{\text{test}}^{*(\zeta)}$. |
| $\vert D_{\text{test}}^{*(\zeta)}\vert $ | Number of participant-wave observations in bootstrap test dataset $D_{\text{test}}^{*(\zeta)}$. |
| $\mathbf{y}_{\text{test}}^{*(s,\zeta)}$ | Outcome vector for substance $s$ in bootstrap test dataset $D_{\text{test}}^{*(\zeta)}$. |
| $\hat{\mathbf{p}}_{\zeta,\text{stack}}^{(s,\eta)}$ | Predicted-probability vector produced by the complete fitted stacked ensemble in bootstrap replicate $\zeta$. |
| $AP_{\zeta, \text{stack}}^{(s, \eta)}$ | Baseline PR-AUC of the complete fitted stacked ensemble in bootstrap replicate $\zeta$. |
| $AP_{j\zeta, \text{stack}}^{\pi(s, \eta)}$ | PR-AUC of the complete fitted stacked ensemble after permuting feature $j$ in bootstrap replicate $\zeta$. |
| $\Delta_{j\zeta}^{(s, \eta)}$ | Signed stacked-model PR-AUC decrease after permutation of feature $j$ in bootstrap replicate $\zeta$. |
| $\bar{\Delta}_j^{(s, \eta)}$ | Mean stacked-model PR-AUC decrease for feature $j$ across $B$ bootstrap replicates. |
| $Q_{0.025}(\cdot)$ | Empirical 2.5th percentile of a bootstrap stacked-permutation importance distribution. |
| $Q_{0.975}(\cdot)$ | Empirical 97.5th percentile of a bootstrap stacked-permutation importance distribution. |
| $CI_{95\%, j}^{(s, \eta)}$ | Percentile-based 95% bootstrap interval for the stacked-model PR-AUC decrease of feature $j$. |
| $CI_{L, j}^{(s, \eta)}$ | Lower bound of $CI_{95\%, j}^{(s, \eta)}$. |
| $\text{Stable}_j^{(s, \eta)}$ | Indicator equal to 1 when $CI_{L, j}^{(s, \eta)} > 0$, indicating stable positive predictive relevance. |

*Note. CS denotes the cross-sectional stacked ensemble and LAG denotes the lagged stacked ensemble. The non-temporal, non-lagged substantive analysis is obtained by filtering the full lagged-model importance ranking; no separate reduced model is fitted.*

### 3.2 Phase 2: Feature Screening and Selection

Building on the datasets constructed in the previous phase, Phase 2 operates on three substance-specific longitudinal panels, one for each substance: alcohol, nicotine, and marijuana. At this stage, no train/test partitioning has been applied, and the full 323-predictor feature space remains unscreened. Phase 2 partitions these datasets safely and identifies the most informative predictors through a training-only screening pipeline. The curated feature matrices produced in this phase serve as input to the ensemble models in Phase 3.

#### 3.2.1 Group-Safe Train/Test Split

Because the dataset contains repeated observations from the same participant, train/test splitting was performed at the participant level rather than the row level. Specifically, 80% of participants were randomly assigned to training and the remaining 20% were reserved for testing, using a group-safe split to prevent information leakage (Kapoor & Narayanan, 2023).

Let:

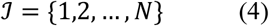

denote the full set of participant indices, as defined in Table 1, where *N* is the number of participants in the analytic dataset. Let:

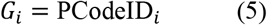

be the participant identifier for individual *i*. The full participant set *ℐ* was partitioned into disjoint training and test participant sets:

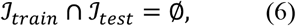

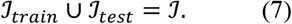

were the corresponding datasets *D*_*train*_ and *D*_*test*_ denote the training and test sets respectively:

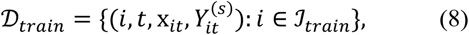

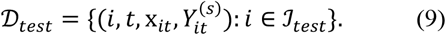

This means all observations from a given participant were assigned entirely to either training or test data. The same participant-level separation was enforced during internal cross-validation, model fitting, feature screening, and threshold calibration, ensuring that every downstream learning step was based only on training participants.

#### 3.2.2 Model Preprocessing

After the group-safe split, preprocessing was performed separately within the training and test partitions. Predictors were classified into numeric and categorical variables. Numeric variables were imputed using median imputation fitted on the training set (Shadbahr et al., 2023). Categorical variables were also imputed and encoded into numeric form. Some models require scaled inputs, including the linear SVM, logistic regression meta-learner, and calibrated multilayer perceptron. For these, standardization was applied within the corresponding pipeline (Shadbahr et al., 2023). Let *T* denote the preprocessing transformation learned from the training data. Then:

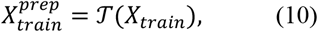

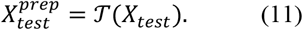

Importantly, *T* was fit using training data only and then applied unchanged to the test set. This ensures that imputation, encoding, and scaling do not use information from held-out participants. Once predictors are transformed into a consistent numeric modeling space, the base learners can be trained.

#### 3.2.3 XGBoost Screening Model

For each substance *s*, an initial XGBoost screening model 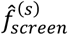, was trained on the preprocessed training data:

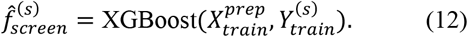

XGBoost (Extreme Gradient Boosting) is a tree-based ensemble method well suited for high-dimensional non-linear tabular data (Chen & Guestrin, 2016). The screening model estimates 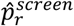, defined as the predicted probability that observation *r*, corresponding to participant *i* at wave *t*, belongs to the positive class:

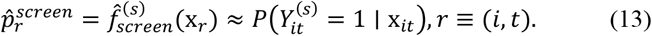

Here, 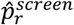 was used exclusively for feature screening and does not represent the final stacked ensemble output. The XGBoost settings were manually specified for training-only feature screening rather than final predictive optimization. A maximum depth of 5 was chosen to allow moderate nonlinear and interaction structure while limiting model complexity. This was treated as a regularization choice rather than a tuned optimal value. A second XGBoost model was used for interaction screening with the same settings. The maximum depth was reduced to 4, since explicit pairwise interaction terms had already been added to the feature space. To account for class imbalance, positive cases were assigned a higher weight during training (Buda et al., 2018). The weight ratio was derived from the class distribution in the training data. All models were implemented using the XGBoost Python library (Chen et al., 2024). For more details, full hyperparameter specifications are provided in Table A1 in Appendix A. No hyperparameter grid search was performed at this stage, as feature screening and model optimization represent distinct pipeline steps (Pudjihartono et al., 2022).

#### 3.2.4 Permutation Importance for Feature Ranking

Feature importance was computed using permutation importance. For each feature *j*, its values were randomly shuffled across observations while all other features were held fixed. The importance score was defined as the decrease in PR-AUC after shuffling:

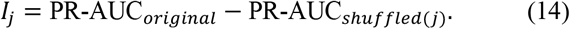

where PR-AUC_*original*_ is the baseline test-set PR-AUC before permutation and PR-AUC_*shuffled(j)*_ is the PR-AUC after randomly permuting feature *j*. The area under the precision-recall curve, or PR-AUC, reflects how well a model detects positive cases across different classification thresholds. In imbalanced settings, this is more meaningful than ROC-AUC because it is sensitive to how the model performs on the minority class specifically, rather than across both classes equally (Williams, 2021). Since substance use outcomes in this dataset were relatively rare, PR-AUC provided a more informative basis for ranking feature importance.

A larger *I*_*j*_indicates that feature *j* contributes more substantially to model performance. Permutation importance was selected over built-in XGBoost measures, such as gain, weight, or cover because those metrics summarize how features are used internally within the tree ensemble. They may not directly reflect how much a feature contributes to out-of-sample classification performance (Fisher et al., 2019).

The importance scores were then sorted:

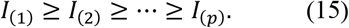

This ranked distribution serves as the basis for feature tiering in the next steps.

#### 3.2.5 Feature Tiering into Strong, Moderate, and Weak Predictors

Features were categorized into tiers using fixed cut points on the permutation-importance scores from Equation (15) (Breiman, 2001; Fisher et al., 2019). The cut points were derived from the alcohol importance distribution curve, which had the most balanced class ratio among the three substances. Feature importance estimates are sensitive to class imbalance, and more balanced distributions produce more stable rankings (Dube & Verster, 2023; Tharwat, 2021). The same thresholds were then applied consistently across all three substances to ensure comparability of retained feature sets.

Specifically:

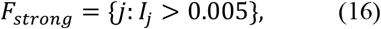

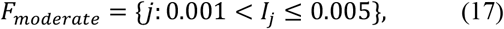

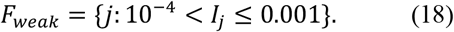

Features with:

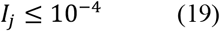

were not retained for tier-based carry-forward.

Each threshold targeted a distinct segment of the permutation-importance distribution. Features above 0.005 formed a small, clearly separable high-impact group. They aligned with a natural inflection point in the sorted importance curve, producing a visible reduction in PR-AUC when permuted. The threshold of 0.001 separated moderate predictors from the long tail of lower-magnitude effects. The threshold 10^−4^ served as a lower noise-floor cutoff, retaining weak but nonzero predictors while excluding features whose permutation impact was effectively negligible.

Thus, the tiering rule can be summarized as:

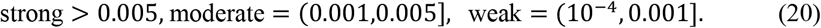

Strong and moderate features were carried forward as main-effect candidates. Weak features were not retained as main effects but were considered in the subsequent interaction-screening stage, as they may still contribute through interactions with stronger predictors.

#### 3.2.6 SHAP Interpretation of the Screening Model

After the XGBoost screening model was trained and permutation importance was computed, SHAP (SHapley Additive exPlanations) values were generated to support model interpretation (Lundberg & Lee, 2017). SHAP was used to explain how individual features contributed to model predictions, but not as the primary criterion for feature tiering.

For observation *r*, the SHAP decomposition is:

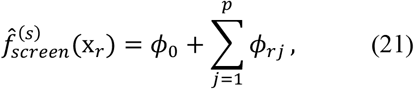

where *ϕ*_0_ is the baseline model output and *ϕ*_*rj*_ is the contribution of feature *j*for observation *r*.

SHAP was included to complement permutation importance, which does not indicate the direction or observation-level contribution of individual features (Fisher et al., 2019; Lundberg & Lee, 2017). In contrast, SHAP decomposes each prediction into feature-level contributions, supporting the interpretation and identification of which features increase or decrease predicted risk across participants

#### 3.2.7 Interaction Generation and Screening

Some variables with small marginal importance may still contribute through interactions (see Section 3.2.5). For this reason, top weak features were retained for interaction screening against strong and moderate predictors. Candidate pairwise interaction terms were defined as:

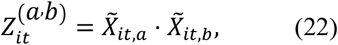

where 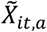 and 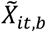 represent encoded or transformed predictors after preprocessing. Interactions were reconstructed after preprocessing and encoding so that they corresponded to the transformed feature space used by the models.

Interaction importance was assessed using the same permutation importance framework described in Section 3.2.4, where *I*_*ab*_ denotes the drop in PR-AUC when interaction term (*a, b*) was shuffled:

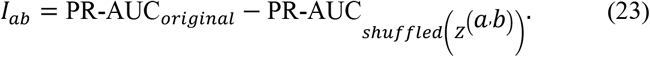

Only retained interaction terms and their source variables were carried forward into the final modeling feature set. This step allowed weak predictors to enter the model when they contributed meaningfully through joint effects with stronger predictors.

#### 3.2.8 Final Feature Set Construction

The final selected feature set combined strong predictors, moderate predictors, and source variables needed to reconstruct retained interactions:

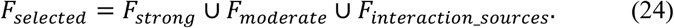

The selected feature set was learned from the training data only. It was then applied unchanged to both training and test matrices. Let:

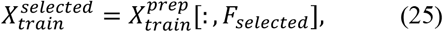

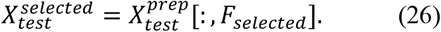

denote the final training and test matrices restricted to selected features. This step ensured that the test set did not influence feature selection while preserving identical feature spaces for model training and evaluation. The resulting selected matrices were used as the input for the cross-sectional and lagged stacking model.

#### 3.2.9 Phase 2: Output and Transition

Sections 3.2.1 through 3.2.8 produced a final selected feature matrix for each of the three substances. This matrix combined strong and moderate main-effect predictors retained through permutation importance tiering with the source variables needed to reconstruct pairwise interaction terms. All selection decisions were fit on training data only and applied unchanged to the test set. This ensured the test set remained entirely unseen at this point, allowing Phase 3 to produce unbiased performance estimates. These feature matrices serve as the direct input to Phase 3. There, two parallel stacked ensemble models are trained: one using current-wave features only, and one augmented with lagged temporal information.

### 3.3 Phase 3: Cross-Sectional and Lagged Stacked Models

Phase 3 built directly on the output of Phase 2, using the final selected feature matrices as its starting point. Two parallel stacked ensemble models were constructed, both sharing the same six-learner architecture but differing in their input feature sets. The cross-sectional stack used only current-wave selected predictors and retained interactions. The lagged stack augmented those same features with prior-wave predictors, developmental timing, and substance-use history variables.

#### 3.3.1 Cross-Sectional Stacking Model

The cross-sectional stacking model used selected current-wave predictors, wave position (defined in Section 3.3.3), and retained interactions, but excluded lagged predictors and prior substance-history features. This configuration served as a cross-sectional benchmark against which the added value of temporal information could later be evaluated.

Let:

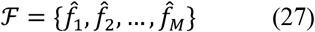

denote the set of *M* base learners. In this study, the stacking ensemble included six base learners: a calibrated linear support vector machine, histogram-based gradient boosting, balanced random forest, LightGBM, XGBoost, and a calibrated multilayer perceptron.

The base learners were selected to provide complementary modeling strengths. Full hyperparameter specifications for all base learners and the meta-learner are provided in Table A2 and A3 in Appendix A. The calibrated linear SVM captured approximately linear decision boundaries in a high-dimensional feature space while producing calibrated probabilities for stacking (Silva Filho et al., 2023). Histogram-based gradient boosting and LightGBM were included as scalable tree-based learners. They use discretized feature bins and leaf-wise tree growth to efficiently model nonlinear relationships and higher-order interactions in high-dimensional tabular data (Ke et al., 2017). XGBoost, as described in Section 3.2.3, provided an additional tree-based learner with regularized boosting. Though all three are tree-based, their differences in feature partitioning, interaction handling, and regularization strategies ensured complementary contributions. Balanced random forest addresses class imbalance through balanced bootstrap sampling while also capturing nonlinear feature relationships (Lemaître et al., 2017). Finally, the calibrated multilayer perceptron provided a neural-network-based learner capable of modeling flexible nonlinear patterns distinct from tree-based approaches (Sarker, 2021).

Together, these learners increased ensemble diversity, allowing the Elastic Net meta-learner to combine linear, tree-based, boosting-based, imbalance-aware, and neural predictive signals (Mienye & Sun, 2022). Potential redundancy among these learners was further controlled through its regularization of correlated base-model predictions. This allowed the final model to emphasize only the learners that improved generalization.

Each base learner produced a predicted probability of substance use for observation *r* from base learner *m*:

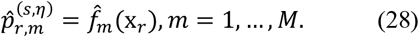

To avoid overfitting in the meta-learner, out-of-fold predictions were generated using group-aware cross-validation (Allgaier & Pryss, 2024; Naimi & Balzer, 2018). For fold *q*, the base model was trained on all other folds:

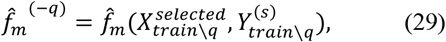

and used to predict observations in fold *q*:

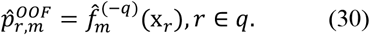

These predictions form the base probability matrix:

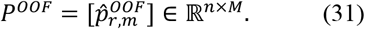

The out-of-fold structure ensures that each training prediction supplied to the meta-learner is generated by a model that did not train on that observation. This design reduces the risk of overfitting and encourages redundancy generalization across unseen participant groups.

#### 3.3.2 Meta-Feature Construction and Elastic Net Meta-Learner

The meta-feature vector *z*_*r*_ was constructed from base-learner predictions and included raw probabilities, logit-transformed probabilities, and pairwise products. Each component was designed to capture different aspects of base-learner behavior.

Raw probabilities represent each base learner’s direct estimate of substance-use risk for a participant-wave observation. Logit-transformed probabilities place those estimates on an odds scale. This is compatible with the Elastic Net logistic regression meta-learner and helps distinguish observations at the extremes of predicted risk (Zian et al., 2021). Pairwise products between base-learner predictions were included to capture agreement across different model families (Dey & Mathur, 2023). When both a tree-based model and a linear model assign high risk to the same adolescent-wave observation, their product becomes large. This indicates that the signal is supported by multiple modeling perspectives.

Together, these meta-features allow the Elastic Net meta-learner to combine individual probability estimates with information about model confidence and cross-learner agreement. The logit transformation was defined as:

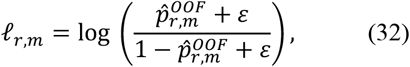

Pairwise probability products were defined as:

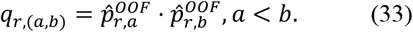

The meta-learner was an Elastic Net logistic regression that estimates the probability of substance use from the meta-feature vector:

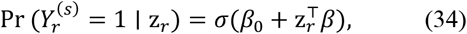

where *σ*(*u*) denotes the sigmoid function:

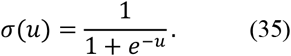

The Elastic Net objective minimizes the regularized binary cross-entropy loss. It drives the meta-learner to assign high probability to true positive cases and low probability to true negative cases.

Specifically, the loss function was defined as:

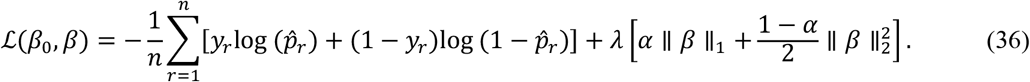

The final stacked prediction is the estimated probability of substance use for observation *r*:

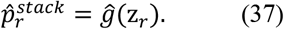

This meta-learning step strengthened the identification of adolescents at elevated risk by leveraging the strengths of all six base learners collectively (Tay et al., 2023). The use of Elastic Net regularization was appropriate because the meta-feature space contains correlated quantities derived from the same base predictions. The L1 penalty shrinks less informative meta-features toward zero, while the L2 penalty stabilizes estimates when meta-features are correlated, reducing overfitting and improving generalization (Zou & Hastie, 2005). The model-specific importance scores, metalearner coefficients, and model-weighted importance measures derived from this ensemble are formally defined in Section 3.5.1.

#### 3.3.3 Longitudinal Feature Engineering

Adolescent substance use follows longitudinal trajectories shaped by prior risk-factor exposure and behavioral history. Temporal context is therefore essential for capturing the developmental nature of risk (Marceau, 2023; Rodríguez-Ruiz et al., 2021). To incorporate this temporal context, the lagged pipeline expanded the cross-sectional feature set with prior-wave predictors and substance-history summaries. Each temporal measure was included to capture a different aspect of adolescent risk over time as summarized in Table 2.

**Table 2.** Temporal and Prior-History Features Used in the Lagged Model. *Each row describes one lagged or prior-history feature included in the LAG model*

| Feature group | Notation | Description | Question answered | What it captures |
| --- | --- | --- | --- | --- |
| **Lagged predictors** | $X_{it,j}^{(\ell)} = X_{i,t-\ell,j}$ | Prior observed values of selected predictors up to four previous assessments. | What were this adolescent's previous risk-factor values? | Earlier demographic/contextual, psychosocial, behavioral, family, peer, school, or physical-health conditions. |
| **Wave position** | $w_t$ | Numeric wave indicator. | Where is this adolescent in developmental time? | Developmental timing and progression across adolescence. |
| **Prior-use indicators** | $\text{prev}_{it}^{(s,k)} = Y_{i,t-k}^{(s)}$ | Substance-use status at prior observed assessments. | Did the adolescent use the substance recently? | Recent use and continuation risk. |
| **Cumulative prior use** | $\text{cumcount}_{it}^{(s)}$ | Count of prior observed assessments with substance use. | How many prior observed assessments showed substance use? | Accumulated exposure or repeated prior use. |
| **Prior assessment count** | $\text{priorwavecount}_{it}$ | Number of prior observed assessments available. | How much prior information is available for this adolescent? | Amount of historical information available for each participant. |
| **Prior-use proportion** | $\text{cumprop}_{it}^{(s)}$ | Proportion of prior observed assessments with substance use. | How consistently has the adolescent used across available history? | Persistence of use relative to available history. |
| **Ever-prior use** | $\text{everprior}_{it}^{(s)}$ | Indicator for any prior observed substance use. | Has the adolescent ever used the substance before? | Whether initiation had already occurred before the current wave. |

*Note.* This table summarizes all temporal features, their notation, and what each one captures in the context of adolescent substance-use risk prediction. Lagged predictors and prior-use history variables are constructed only from observed prior assessments; missing prior waves do not contribute.

For each selected raw/source predictor *j*, lagged values up to four prior observed assessments were created. This window was selected to capture the most developmentally proximal prior exposures while avoiding the inclusion of temporally distant observations that may be less predictive of current risk (Hawes et al., 2023):

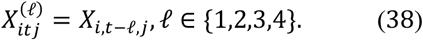

where 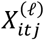 represents the value of predictor *j* for participant *i* at lag *ℓ* relative to wave *t*. These lagged predictors capture whether earlier conditions are associated with later use. For example, a prior-wave increase in peer deviance, emotional distress, or reduced parental monitoring may carry predictive information even if the current-wave value alone is insufficient.

Developmental timing was represented by the wave-position feature *w*_*t*_, first introduced in Section 3.3.1 as a numeric indicator of the adolescent’s location in the study timeline:

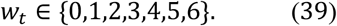

Wave position was preferred over chronological age because participants in the ABCD study were recruited within a narrow age band. Wave number is therefore a more precise indicator of developmental progression. It captures assessment-specific effects that age alone may not reflect (Garavan et al., 2018).

Prior-use indicators were defined as:

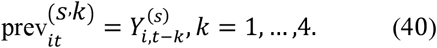

where 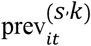 captures prior history of the same substance at each of the four most recent observed assessments. They are important because recent and earlier prior use may reflect behavioral persistence or continuation risk (Sullivan et al., 2022).

The cumulative count of prior observed use was:

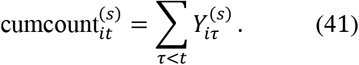

where 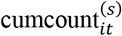 measures how many previous observed assessments showed evidence of use before the current visit (Nasir et al., 2021).

The number of observed prior assessments was:

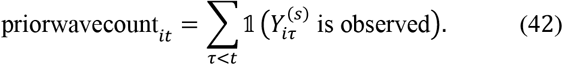

where priorwavecount_*it*_captures how much prior information is available for participant *i* at wave *t*. It prevents cumulative counts from being interpreted without accounting the amount of follow-up available (Wei et al., 2026). The cumulative prior-use proportion was:

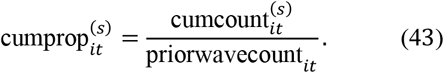

where 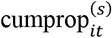 represents the proportion of previously observed assessments in which substance use was recorded (Atias et al., 2024). It is useful because two adolescents may both have two prior uses, but one may have two uses across two observed assessments while another may have two uses across five.

Finally, any prior observed use was represented as:

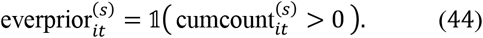

where 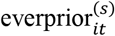 distinguishes adolescents with any prior evidence of use from those with no prior evidence before wave *t*. It is important because the risk process may differ between first-time initiation and continuation after prior use.

Observations were included in the lagged model only if the participant had at least one prior observed assessment, ensuring that at least one lag value could be constructed. Lag features were therefore constructed from prior observed waves within each participant’s available record. Complete participation at every scheduled wave was not required.

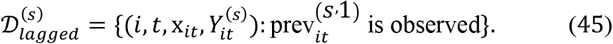

where 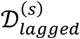 is the filtered dataset that ensured that lagged models were based on observations with valid prior-history information. First-observed rows for each participant were excluded from the lagged model because no previous observation was available to construct lag-1 features or prior-use indicators.

#### 3.3.4 Lagged Stacking Model

After constructing the longitudinal feature space in Section 3.3.3, the same stacked-ensemble architecture described in Sections 3.3.1 and 3.3.2 was applied to the lagged dataset. This step evaluated whether adding temporal information improved prediction beyond the cross-sectional model.

The lagged stack used the same six base learner families as the cross-sectional stack, but the input vector was expanded to include both current and historical information:

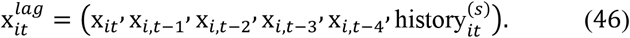

where 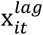 represents the expanded input vector for participant *i* at wave *t*, combining current-wave predictors with lagged and prior-use history features. Here 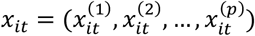 contains the current-wave selected predictors, 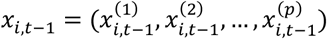 through 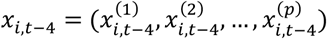 contain the values of those same predictors from the four most recent prior observed assessments, and 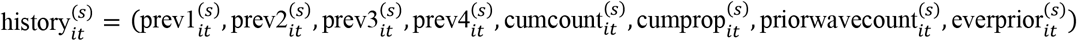 contains the prior-use summaries defined in Section 3.3.3.

Each base learner produced a predicted probability using the expanded lagged feature vector:

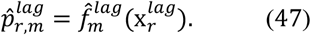

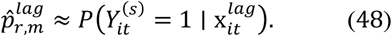

where *r* ≡ (*i, t*), so each prediction corresponds to one participant at one observed wave. Unlike the cross-sectional prediction in Section 3.3.1, which used only current-wave selected predictors, 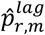 is based on the adolescent’s current profile, prior risk-factor values, developmental timing, and substance-use history. As in the cross-sectional stack, group-aware out-of-fold predictions were generated for each base learner and passed into the Elastic Net meta-learner.

The final lagged stacked prediction was:

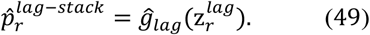

where 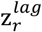 contains meta-features derived from the lagged base-learner predictions, following the same meta-feature construction described in Section 3.3.2. The group-aware validation strategy from Section 3.2.1 was maintained to ensure participant-level separation between training and validation folds. This prevented overlap when generating predictions for the meta-learner.

The final lagged stacking model can therefore be interpreted as:

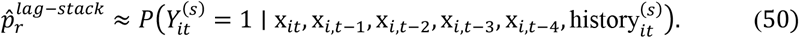

where 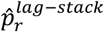 is the estimated probability of substance use for observation *r* using both current and historical features. Because the lagged and cross-sectional models used the same selected feature foundation and ensemble architecture, any differences in performance can be attributed to the added contribution of temporal information rather than changes in the modeling framework.

#### 3.3.5 Phase 3: Output and Transition

Sections 3.3.1 through 3.3.4 produced two substance-specific stacked probability estimates per observation, one from the cross-sectional stack and one from the lagged stack. These estimates served as the direct input to Phase 4, which evaluates model performance on the held-out test set.

### 3.4 Phase 4: Model Evaluation

Phase 4 received as input the stacked probability estimates produced by both the cross-sectional and lagged models in Phase 3. It also received the out-of-fold training predictions used for threshold calibration. The model architecture, feature set, and meta-learner coefficients were fully fixed, and no further training or feature selection was performed. A recall-constrained threshold of 0.80 was applied to convert continuous risk scores into binary classifications, consistent with the prevention-oriented screening goal of the study. Final model performance was then evaluated on the held-out test set.

#### 3.4.1 Threshold Calibration and Final Evaluation

The probabilistic outputs of the cross-sectional and lagged stacked models were converted into binary classifications through threshold calibration. For any candidate threshold *c*, predicted probabilities were converted into binary class labels as follows:

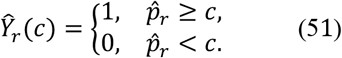

Here, 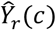 denotes the predicted class label for observation *r*at threshold *c*, where 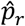 is the stacked model’s predicted probability of substance use. Different thresholds produce different tradeoffs between sensitivity and false positives. A lower threshold generally increases recall but may reduce precision by increasing false positives. A higher threshold has the opposite effect.

Because this study targets prevention-oriented screening, a minimum recall of 0.80 was used as the sensitivity benchmark (Patnode et al., 2020). This reflects the premise that missing a true high-risk case carries greater cost than a false positive. The optimal threshold *c*\* was selected to maximize precision subject to this recall constraint:

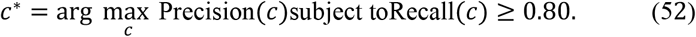

Recall was defined as:

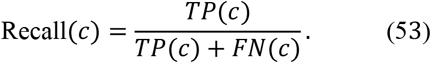

where *TP*(*c*) denotes true positives and *FN*(*c*) denotes false negatives at threshold *c*. Recall was prioritized because false negatives correspond to adolescents with substance-use evidence who were not identified by the model. Precision was defined as:

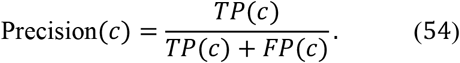

where *FP*(*c*) denotes false positives at threshold *c*. Together, equations (52) to (54) formalize the screening-oriented decision rule. The model first satisfies a minimum sensitivity requirement and then selects the threshold with the best positive predictive value under that constraint.

Threshold selection was performed using training out-of-fold predictions rather than the held-out test set, following the same leakage-prevention strategy described in Section 3.3.2.

Once *c*\* was selected, it was applied unchanged to the held-out test predictions:

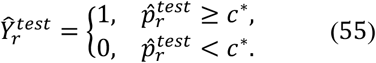

where 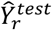 test represents the final binary classification for observation *r* on the held-out test set. The test set was used only after the model, feature set, meta-learner, and threshold had already been fixed, preserving it as an unbiased estimate of generalization to unseen adolescents.

Final model performance was evaluated using both threshold-independent and threshold-dependent metrics. Threshold-independent metrics included ROC-AUC and PR-AUC, which evaluate the ranking quality of predicted probabilities across possible thresholds (Williams, 2021). Threshold-dependent metrics included the confusion matrix, precision, recall, and top-risk stratification metrics. Precision at the top *q*%of predicted risk was defined as:

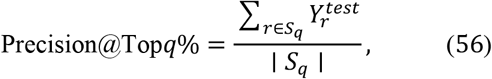

where *S*_*q*_ denotes the set of test observations ranked in the top *q*% by predicted probability. This metric is especially relevant for prevention and resource allocation because interventions may be targeted to a limited high-risk subgroup rather than the entire population (Green et al., 2024; Rakers et al., 2024) I think the sentence of 3.4.2 is good.

#### 3.4.2 Phase 4: Output and Transition

Phase 4 produced a set of final performance metrics for both the cross-sectional and lagged stacked models across the three substances. These covered threshold-independent ranking metrics, threshold-dependent classification metrics, and top-risk stratification measures. These results served as the direct input to Phase 5.

### 3.5 Phase 5: Model Interpretation

Phase 5 received as input the fitted stacked models, meta-learner coefficients, and held-out test predictions produced in Phases 3 and 4. Building on the performance evaluation completed in Phase 4, this phase shifted focus from predictive accuracy to model interpretation, examining which features drove substance-use risk predictions, how those contributions varied across substances, and whether the observed patterns were stable across resampled participants.

#### 3.5.1 Base-Learner Permutation Importance

For each fitted stacked ensemble, permutation importance was computed separately for each of the six base learners included in ℳ: calibrated Linear Support Vector Machine (SVM), HistGradientBoosting (HGB), Balanced Random Forest (BRF), LightGBM (LGBM), XGBoost (XGB), and a calibrated Multilayer Perceptron (MLP). PR-AUC-based permutation importance was selected because it is model-agnostic and can be applied uniformly across all six base learners without requiring model-specific explanation procedures. This provided a consistent performance-based importance metric across all six heterogeneous base learners, substance-specific outcomes, and model designs (Fisher et al., 2019).

Let *s, η, m* and *j* index the substance-specific outcome, model design, base learner, and feature, respectively, as defined in Table 1. Here, ℳ = 6 and 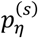 are as defined in Table 1.

For each fitted base learner *m*, the baseline test-set PR-AUC was defined as:

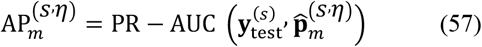

where 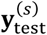 is the observed test-set outcome vector for substance *s*, and 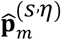 is the predicted-probability vector produced by fitted base learner 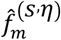

To estimate the predictive relevance of feature *j*, the permutation operation *π*_*j*_randomly reordered the values of that feature in the held-out test set while leaving all remaining predictors unchanged. For permutation repeat *ν* = 1, …, *R*_*π*_, where *R*_*π*_ = 5, the resulting PR-AUC was defined as:

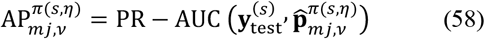

where 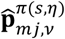 denotes the predicted-probability vector produced by base learner *m*after feature *j* was permuted during repeat *ν*.

The model-specific permutation importance of feature *j* for base learner *m*was calculated as the absolute mean decrease in PR-AUC:

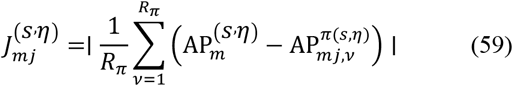

The absolute value was used at the ranking stage so that predictors associated with meaningful changes in predictive performance were retained. This ensured retention even when empirical permutation variability produced small negative PR-AUC differences. Because importance magnitudes can differ across learning algorithms, the permutation-importance scores were then normalized within each base learner before aggregation:

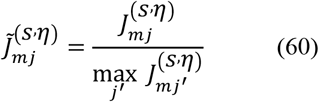

where 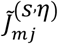 represents the normalized importance of feature *j* for base learner *m*, scaled relative to the maximum importance observed across all features for that learner in substance *s* under model design η. This transformation placed each base learner’s feature-importance scores on a relative common scale. When all permutation-importance scores for a base learner were zero, its normalized importance vector remained zero.

#### 3.5.2 Meta-Learner-Weighted Beta-Aggregated Feature Importance

The stacked ensembles combined the outputs of the six base learners using an Elastic Net Logistic Regression meta-learner *ĝ*^(*s, η*)^. Stacked generalization integrates predictions from multiple first-level learners through a second-level learner trained on their outputs (Naimi & Balzer, 2018; Nguemkam Tebou et al., 2026). Elastic Net regularization combines *L*_1_and *L*_2_penalties, permitting coefficient shrinkage while retaining information among correlated meta-features (Zou & Hastie, 2005).

The meta-feature vector, 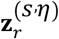, was constructed as described in Section 3.3.2. Model-level weights were then derived by beta-aggregating normalized permutation-importance scores using selected coefficients from the fitted Elastic Net meta-learner.

For model-level importance aggregation, the coefficient block associated with the six raw base-learner probability outputs was examined first. Let 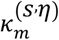 denote the absolute applicable Elastic Net coefficient representing the direct meta-feature contribution of base learner *m*. When all coefficients associated with the six raw probability outputs were zero due to Elastic Net shrinkage, the logit-transformed output coefficients were used instead. If both direct coefficient blocks were entirely zero, equal weights were assigned to the six base learners.

The normalized coefficient-based weight assigned to base learner *m* was defined as:

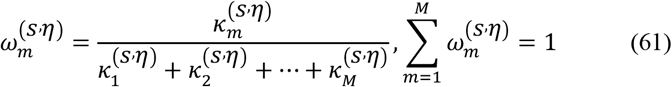

The beta-aggregated permutation importance of feature *j* was then calculated as:

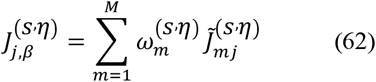

where 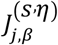 represents the beta-aggregated importance of feature *j* for substance *s* under model design η, producing one ensemble-informed feature ranking per substance and model design. For descriptive reporting, the percentage contribution of each feature was calculated as:

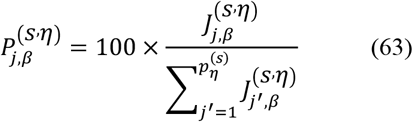

where 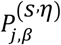 is the percentage contribution of feature *j* to total ensemble importance for substance *s* under model design η. The top *K* = 20 features from each complete model ranking, ordered by descending 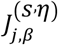, were carried forward to the bootstrap stability analysis.

#### 3.5.3 Cross-Sectional Feature-Importance Analysis

For the cross-sectional stacked ensembles, the beta-aggregated permutation-importance framework was applied to the final encoded feature space for each substance-specific model. This feature space included selected current-wave predictors, reconstructed current-wave interaction terms, and the developmental timing variable (wave number).

For each cross-sectional outcome, permutation importance was computed for all six fitted base learners using the PR-AUC-based procedure defined in Section 3.5.1. These scores were normalized within base learner and combined using the meta-learner weighting procedure defined in Section 3.5.2. The resulting rankings identified the current-wave signals most relevant to predictive performance. These models excluded prior-wave predictor values, prior-use indicators, and cumulative prior-use history measures. This cross-sectional interpretation provided a baseline representation of feature relevance based on predictive signals observed at the current participant-wave observation.

#### 3.5.4 Full Lagged Feature-Importance Analysis

For the lagged stacked ensembles, the same beta-aggregated permutation-importance framework was applied to the complete longitudinal feature space, which incorporated longitudinal features organized into three subsets, as defined in equations 65 to 67.

For each substance *s*, the complete lagged feature set was defined as:

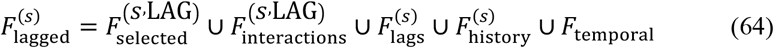

The lagged predictor subset contained prior values of the selected raw and interaction-source predictors:

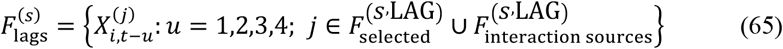

The substance-specific prior-use history subset was defined as:

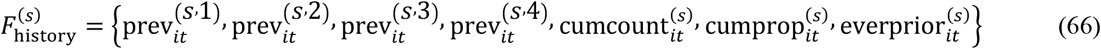

The temporal feature subset was defined as:

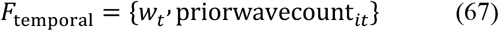

where *w*_*t*_corresponds to the numeric wave-position variable, wave number.

The full lagged ranking quantified the predictive relevance of all information available to the fitted longitudinal ensemble, as defined in Equation 64.

#### 3.5.5 Non-Temporal, Non-Lagged Substantive Feature Analysis

A complementary analysis was conducted for each lagged stacked ensemble to identify substantive current-wave predictors that remained relevant after longitudinal information had been incorporated into the fitted model. This analysis did not involve fitting a separate reduced model. Instead, it began with the beta-aggregated importance ranking obtained from the fitted full lagged ensemble and excluded variables that directly represented prior predictor values, prior substance-use history, or temporal position.

The excluded feature set for substance *s* was defined as:

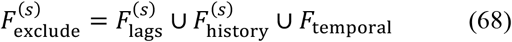

where 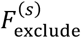 represents the set of features excluded from the substantive analysis, comprising lagged predictors, prior-use history variables, and temporal features.

The retained substantive feature subset was defined as:

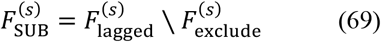

Specifically, 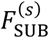 retained current-wave substantive predictors and interaction terms by removing all lagged predictors, prior-use history variables, and temporal features as defined in Equation 68.

Because importance scores were filtered from the fitted full lagged ensemble rather than recalculated from a newly trained reduced model, the importance values reflected the contribution of each retained feature within the complete longitudinal model. This analysis addressed the following interpretive question: which substantive current-wave predictors remained most relevant within a fitted longitudinal ensemble that already had access to prior history?

The percentage contribution of each retained substantive feature was recalculated within the filtered subset:

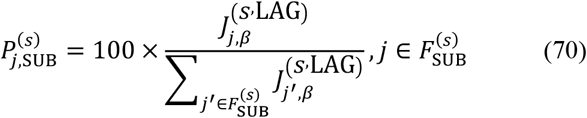

where 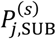 is the percentage contribution of feature *j* to total importance within the substantive feature subset 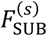 for substance *s*. The top *K* = 20 retained substantive features for each substance were then evaluated through participant-level bootstrap stability analysis.

#### 3.5.6 Participant-Level Bootstrap Stability Analysis of the Complete Stacked Ensemble

Beta-aggregated permutation importance was used to generate the initial feature rankings, whereas uncertainty was assessed using the complete fitted stacked ensemble. This distinction ensured that the reported stability intervals quantified the predictive relevance of selected features in the final ensemble architecture rather than in any individual base learner.

For each analysis type (cross-sectional, full lagged, and lagged substantive), the top *K* = 20 features from the corresponding beta-aggregated ranking were evaluated.

The analytic data included repeated participant-wave observations. Therefore, bootstrap resampling was conducted at the participant level rather than at the individual-row level. This ensured that all repeated observations for each sampled participant remained together within each bootstrap replicate. Cluster-level bootstrapping is appropriate when observations are grouped within higher-level units, such as repeated measurements within participants (Deen & de Rooij, 2020).

A total of *B* = 200 bootstrap replicates were used to ensure stable percentile-based interval estimation (Rousselet et al., 2021). For each replicate *ζ* = 1, …, *B*, participants in the held-out test partition were sampled with replacement:

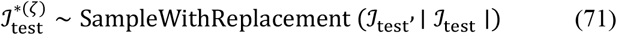

where 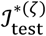 is the bootstrap sample of participant indices drawn with replacement in replicate ζ. All participant-wave observations belonging to the sampled participants were retained to form the bootstrap test dataset, 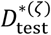.

Within bootstrap replicate *ζ*, the baseline PR-AUC of the complete fitted stacked ensemble was calculated as:

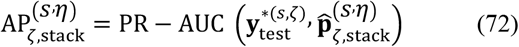

where 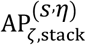 denotes this baseline for substance *s* under model design η. For each selected feature *j*, its values were permuted within 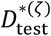. Predictions were then regenerated through the complete fitted ensemble. This involved obtaining outputs from all six base learners, reconstructing the meta-feature representation, and applying the fitted Elastic Net meta-learner.

The stacked-model permutation importance of feature *j* in bootstrap replicate *ζ*was defined as:

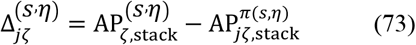

where 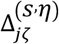 captures the signed PR-AUC difference after permuting feature *j*. Unlike the initial beta-aggregated ranking, which used absolute mean decreases in base-learner PR-AUC, the bootstrap analysis retained the signed stacked-model PR-AUC difference.

The bootstrap mean importance was computed as:

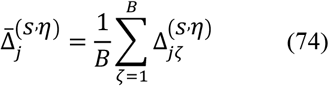

where 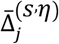 reflects the mean importance of feature *j* across all *B* replicates.

Percentile-based 95% bootstrap intervals were obtained from the empirical distribution of 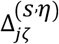 across the *B* participant-level resamples (Rousselet et al., 2021):

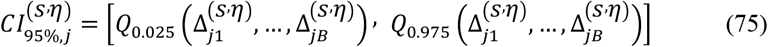

where 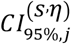 refers to the 95% stability interval for the stacked-model permutation importance of feature *j*. A feature was classified as exhibiting stable positive predictive relevance when the lower bound of its bootstrap interval exceeded zero:

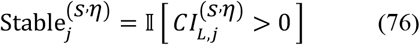

This criterion indicates that permutation of the feature consistently reduced stacked-ensemble PR-AUC across participant-level bootstrap resamples. It should not be interpreted as evidence of a causal relationship or as indicating whether the feature increased or decreased predicted substance-use risk.

#### 5.5.7 Phase 5: Output

Phase 5 generated three complementary beta-aggregated feature-importance rankings for each substance-specific outcome. Following the procedure described in Sections 3.5.1 and 3.5.2, PR-AUC-based permutation importance was computed, normalized, and aggregated for each fitted cross-sectional and full lagged ensemble. The lagged substantive ranking was derived as described in Section 3.5.5. The top *K* = 20 features from each ranking were then evaluated for stability. This was done by measuring the signed decrease in PR-AUC of the complete stacked ensemble after feature permutation across *B* = 200 participant-level resamples. The resulting 95% stability intervals were used to identified top-ranked features whose positive predictive relevance was consistent across resampled test participants (Rousselet et al., 2021). Because the full lagged models included a broader feature space than the cross-sectional models, percentage contributions were interpreted within each analysis rather than compared directly across model designs. The resulting rankings and stability intervals are presented in the Results section.

## 4. Results

Results are presented across four sections. The first describes the data source and study sample. The second and third sections present cross-sectional and lagged stacked model performance respectively, evaluating whether longitudinal information improves prediction. The fourth removes temporal variables from the lagged models to identify which non-temporal factors remain most associated with substance-use risk.

### 4.1 Data Source and Study Sample

This study used data from the Adolescent Brain Cognitive Development (ABCD) Study Release 6.0, one of the largest longitudinal studies of adolescent brain and cognitive development in the United States (Hoffman, 2024). The ABCD Study enrolled 11,880 participants at baseline between ages 9 and 10, following them across up to six assessment waves after baseline through age 15 to 16 in Release 6.0. The release includes data from 11,868 participants, reflecting the full cohort minus 12 individuals who withdrew consent to share their data (ABCD Study, 2025).

This study focused exclusively on survey-based data organized across five primary domains: demographics, friends, family and community, mental health, physical health, and substance use. While the ABCD dataset also contains neuroimaging, neurocognitive tasks, biospecimens, wearable sensors, and genetics, these data types were outside the scope of this study (ABCD Study, 2025). Within each domain, variables were drawn from validated questionnaires administered directly to participants. Although parent and caregiver-reported instruments are also available in the ABCD dataset, this study used only youth-reported measures. Each questionnaire typically comprised multiple tables and dozens of individual items. Assessment frequency varied across instruments, with some collected at every annual wave, others every two years, and some every six months.

Variable inclusion was based on missingness across the six available waves. Variables with more than 50% missing data across waves were excluded, as retaining them would have compromised the lagged feature construction that required consistent prior-wave information (Young & Johnson, 2015). The final analytic dataset included 323 variables distributed across domains as follows: 253 from mental health, 48 from friends, family and community, 14 from demographics, and 8 from physical health. A detailed breakdown of variables by domain and subdomain is provided in Table C1 and C2 in the Appendix.

ABCD was selected because its longitudinal design allows substance-use vulnerability to be modeled as a developmental process rather than a static outcome. The combination of repeated assessments, diverse predictor domains, and a large nationally recruited sample makes it particularly well-suited for longitudinal machine learning modeling. These features supported the development and evaluation of substance-use risk prediction models across multiple assessment waves (Hoffman, 2024).

### 4.2 Model Performance and Predictive Validity Across Substances

Tables 3, 4, and 5 report held-out test performance for alcohol, nicotine, and marijuana respectively, with all models ranked by PR-AUC. Three findings emerge from these results.

**Table 3.** Held-out test performance for alcohol initiation prediction across candidate models, ranked by PR-AUC.

| Rank | Model | PR-AUC | ROC-AUC | Recall @ ~0.80 | Precision @ ~0.80 | F1 @ ~0.80 | Accuracy |
| --- | --- | --- | --- | --- | --- | --- | --- |
| 1 | Lagged Stacked Ensemble | 0.598 | 0.854 | 0.817 | 0.361 | 0.500 | 0.741 |
| 2 | XGBoost | 0.463 | 0.786 | 0.801 | 0.295 | 0.431 | 0.640 |
| 3 | HistGradientBoosting | 0.460 | 0.783 | 0.801 | 0.293 | 0.429 | 0.637 |
| 4 | Balanced Random Forest | 0.444 | 0.772 | 0.801 | 0.292 | 0.428 | 0.636 |
| 5 | Cross-Sectional Stack | 0.414 | 0.758 | 0.818 | 0.265 | 0.400 | 0.583 |
| 6 | Logistic Regression | 0.401 | 0.749 | 0.801 | 0.261 | 0.394 | 0.580 |
| 7 | Linear SVM Calibrated | 0.399 | 0.749 | 0.802 | 0.261 | 0.394 | 0.580 |
| 8 | MLP Calibrated | 0.377 | 0.732 | 0.804 | 0.251 | 0.382 | 0.557 |

**Table 4.** Held-out test performance for nicotine initiation prediction across candidate models, ranked by PR-AUC.

| Rank | Model | PR-AUC | ROC-AUC | Recall @ ~0.80 | Precision @ ~0.80 | F1 @ ~0.80 | Accuracy |
| --- | --- | --- | --- | --- | --- | --- | --- |
| 1 | Lagged Stacked Ensemble | 0.548 | 0.917 | 0.813 | 0.245 | 0.376 | 0.869 |
| 2 | HistGradientBoosting | 0.511 | 0.918 | 0.801 | 0.242 | 0.371 | 0.877 |
| 3 | XGBoost | 0.495 | 0.914 | 0.801 | 0.240 | 0.369 | 0.876 |
| 4 | Balanced Random Forest | 0.458 | 0.913 | 0.801 | 0.234 | 0.362 | 0.872 |
| 5 | Linear SVM Calibrated | 0.422 | 0.904 | 0.801 | 0.210 | 0.333 | 0.854 |
| 6 | Logistic Regression | 0.419 | 0.904 | 0.801 | 0.210 | 0.333 | 0.855 |
| 7 | **MLP Calibrated** | 0.395 | 0.883 | 0.809 | 0.175 | 0.288 | 0.819 |
| 8 | **Cross-Sectional Stack** | 0.365 | 0.881 | 0.811 | 0.157 | 0.263 | 0.794 |

**Table 5.**
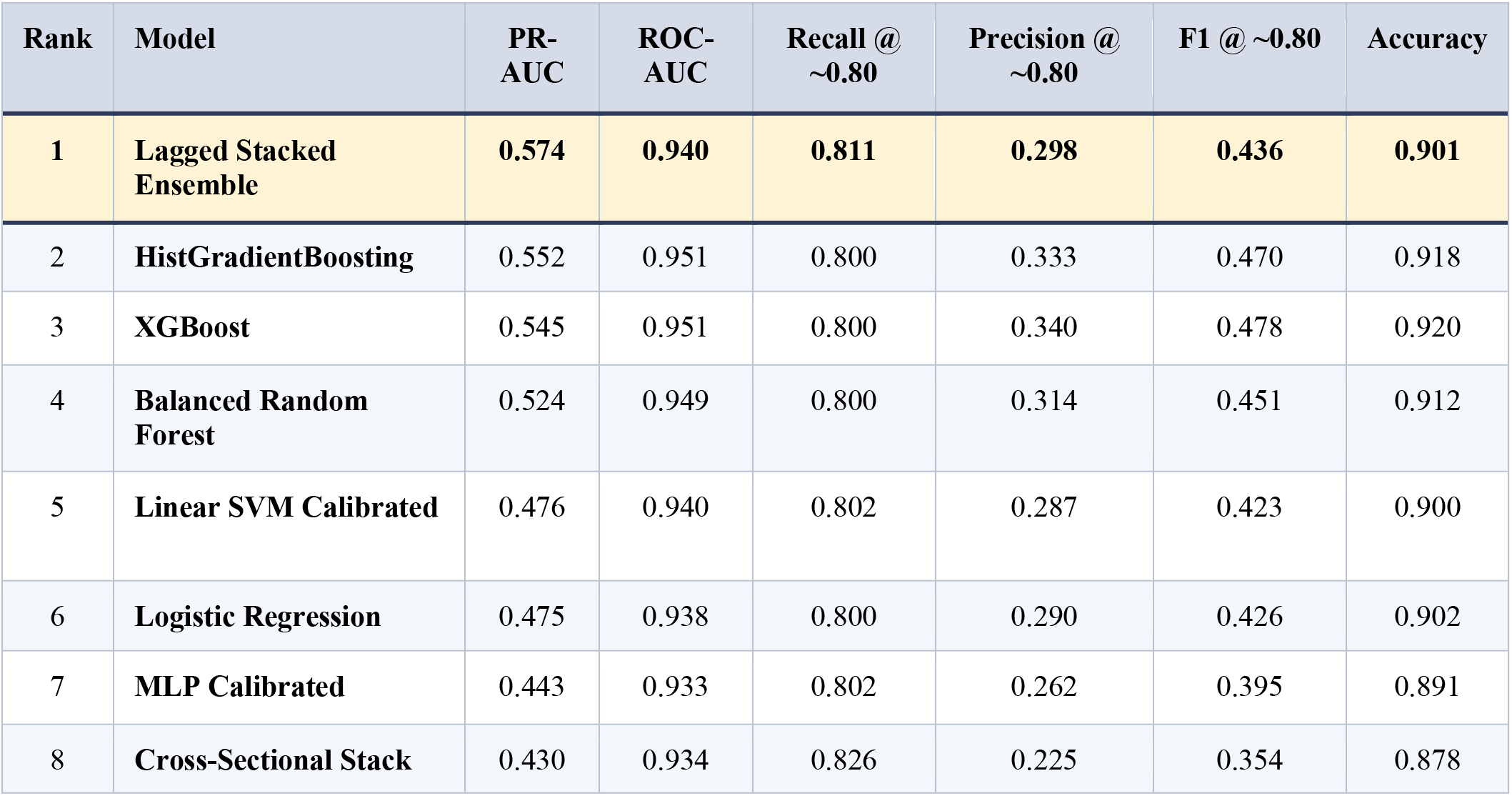
Held-out test performance for marijuana initiation prediction across candidate models, ranked by PR-AUC.

#### 4.2.1 Longitudinal Information Drives Performance Gains

The lagged stacked ensemble achieved the highest PR-AUC across all three substances: 0.598 for alcohol, 0.548 for nicotine, and 0.574 for marijuana. Two comparisons help isolate the source of this advantage. First, the lagged stack substantially outperformed the cross-sectional stack across all three substances. PR-AUC gaps were 0.184 for alcohol, 0.183 for nicotine, and 0.144 for marijuana. Since both stacks share the same ensemble architecture, this difference is attributable to the addition of prior-wave predictors, developmental timing, and substance-use history. This is consistent with prior work showing that lagged predictors capturing prior behavior and developmental trajectories improve risk prediction beyond same-wave information alone (Lauvsnes et al., 2022; Wei et al., 2026).

Second, the lagged stack outperformed all single base learners on PR-AUC across all three substances. For alcohol (Table 3), the best single model was XGBoost (PR-AUC = 0.463), compared to the lagged stack’s 0.598, a gain of 0.135. For nicotine (Table 4), the best single model was HistGradientBoosting (PR-AUC = 0.511), compared to the lagged stack’s 0.548, a gain of 0.037. For marijuana (Table 5), the best single model was HistGradientBoosting (PR-AUC = 0.552), compared to the lagged stack’s 0.574, a gain of 0.022. Notably, the cross-sectional stack ranked last or near last among all models, consistently underperforming single base learners. This suggests that stacking alone does not add predictive value in this setting. The performance advantage of the lagged stack over single models therefore reflects the contribution of longitudinal information rather than ensemble architecture alone (Lauvsnes et al., 2022; Wei et al., 2026).

Together, these two comparisons show that longitudinal information is the primary driver of performance gains in the lagged stack. Stacking without temporal history did not improve over single models, whereas stacking with temporal history did.

#### 4.2.2 Screening Performance at the Recall Threshold

All candidate models achieved recall at or above 0.80 across all three substances. The classification threshold was selected to maximize precision subject to a minimum recall constraint of 0.80. This reflects the prevention-oriented goal of the study, where missing a true high-risk adolescent carries greater cost than a false positive (Patnode et al., 2020). The lagged stacked ensemble achieved recall of 0.817 for alcohol, 0.813 for nicotine, and 0.811 for marijuana. It also achieved the highest precision among models with comparable recall.

Precision was modest across all models, ranging from 0.245 to 0.361. This is expected given the class imbalance, where positive cases represented approximately 18% for alcohol and 5% for nicotine and marijuana (Tharwat, 2021).

These results compare favorably to prior studies with related goals. Kim et al. (2025) predicted substance use across three international cohorts using XGBoost, reporting ROC-AUC values between 0.70 and 0.80 without a recall constraint. The present study achieved higher ROC-AUC values of 0.854, 0.917, and 0.940 for alcohol, nicotine, and marijuana respectively. Rajapaksha et al. (2025) and Wang et al. (2025) predicted disorder-level outcomes among adolescent and young adult substance users, reporting AUC values between 0.64 and 0.75. While their outcome definition differs, both studies share the continuation-oriented framing of the present study, making the comparison more meaningful than initiation-focused designs. Pelham et al. (2024) specifically targeted substance-naïve youth for initiation prediction using only baseline ABCD predictors, reporting AUC values between 0.61 and 0.67. Their lower performance likely reflects the absence of longitudinal history rather than differences in modeling approach. Across these comparisons, the present framework achieved higher discrimination while explicitly targeting a prevention-relevant recall threshold.

#### 4.2.3 Outcome Prevalence Moderates Ensemble Gains

The performance advantage of the lagged stacked ensemble over the best single model decreased substantially as outcome prevalence decreased. For alcohol, the most prevalent outcome at approximately 18% positive cases, the PR-AUC advantage was 0.135. For nicotine and marijuana, both at approximately 5% positive cases, the advantage narrowed to 0.037 and 0.022 respectively. This pattern is not unexpected. With more positive cases, there is more signal available for the meta-learner to integrate across diverse base learners. When the outcome is rare, individual strong learners can approach the ensemble’s performance (Ayodele, 2023; Khan et al., 2023). This finding suggests that stacking provides the largest gains when the outcome is sufficiently prevalent to generate diverse and complementary signals across base learners.

The narrowing advantage for rarer outcomes is perhaps best illustrated by the marijuana results (Table 5), where XGBoost outperformed the lagged stack on F1 despite the stack’s higher PR-AUC. XGBoost achieved a higher F1 score (0.478 vs. 0.436) and higher precision (0.340 vs. 0.298) at a comparable recall level. The lagged stacked ensemble is nonetheless preferred here because PR-AUC summarizes discrimination performance across the full range of operating thresholds, not just at the selected recall point (Kim et al., 2025). The lagged stack’s higher PR-AUC (0.574 vs. 0.545) indicates it ranks positive cases more reliably across all thresholds. In prevention-oriented screening, where the goal is to maximize sensitivity to true high-risk cases, the threshold-free advantage of PR-AUC is the more appropriate criterion for model selection.

### 4.3 Feature Importance Across Modeling Approaches

Feature importance was quantified through a unified score combining importance measures from all six base learners. For each substance, findings are presented as a narrative integrating results across the cross-sectional, full lagged, and non-temporal lagged models.

#### 4.3.1 Alcohol

For alcohol, the distribution of beta-aggregated feature importance changed substantially across modeling approaches. The cross-sectional model was primarily characterized by interaction terms, although Mental Health also contributed the largest share among standalone current-wave domains. Interaction terms accounted for 49.65% of total beta-aggregated importance, with the largest contributions from Friends, Family, and Community × Mental Health interactions (15.76%), Physical Health × Mental Health interactions (10.25%), and Physical Health × Physical Health interactions (9.58%) (Appendix Table B1). At the individual-feature level, wave number was the highest-ranked feature (8.59%), followed by the height-by-weight interaction (7.73%), the interaction between peer shoplifting and relational aggression (6.43%), peer shoplifting as a main effect (6.29%), and the interaction between peer shoplifting and tendency to think carefully before acting (6.13%). Parental knowledge of companions also ranked among the leading current-wave predictors (4.27%). Thus, in the absence of explicitly constructed prior-wave history, alcohol prediction reflected a distributed combination of developmental timing, peer and parenting context, mental-health or behavioral features, physical-health measures, and cross-domain interactions.

The importance profile shifted markedly in the full lagged model. Prior alcohol-use history became the dominant feature area, accounting for 48.93% of total beta-aggregated importance, while temporal features contributed an additional 12.66% (Appendix Table B1). At the individual-feature level, cumulative prior alcohol-use count ranked first (24.63%), followed by alcohol use at the most recent prior wave (19.45%) and wave number (11.06%). Together, these three features accounted for 55.14% of total importance in the full lagged model, compared with 22.75% for the three highest-ranked features in the cross-sectional model. Correspondingly, the contribution of current-wave interaction terms decreased from 49.65% in the cross-sectional model to 16.13% in the full lagged model. These findings indicate that alcohol prediction became concentrated in prior alcohol-use history and temporal information once longitudinal data were included. This contrasts with the broader current-wave feature profile observed in the cross-sectional model.

Physical-health features and their interactions emerged as the dominant retained current-wave predictors once lagged predictors, and temporal and prior-use variables were excluded. Interaction terms again represented the largest feature area, accounting for 59.21% of retained substantive importance. Among interaction categories, Friends, Family, and Community × Physical Health contributed the largest share of retained substantive importance (15.67%), followed by Physical Health × Mental Health (11.89%) and Physical Health × Physical Health (11.45%) (Appendix Table B1). Among individual retained features, weight ranked first (11.41%), followed by the height-by-weight interaction (9.90%), the peer-shoplifting-by-height interaction (9.53%). Positive urgency, reflecting the tendency to act rashly under intense positive emotion, also ranked among the most important retained substantive features. Taken together, although prior alcohol-use history dominated the full lagged model, the remaining current-wave substantive signal was concentrated in physical-health features and their interactions with peer/family context, demographic features, and mental-health or behavioral characteristics. Bootstrap stability analyses confirmed these patterns, with 14, 13, and 11 features showing stable positive predictive relevance in the cross-sectional, full lagged, and lagged substantive analyses, respectively (Appendix B, Figures B1–B3).

#### 4.3.2 Nicotine

For nicotine, the cross-sectional importance profile differed markedly from alcohol, with developmental timing emerging as the primary predictive signal. Wave number alone accounted for 34.68% of total beta-aggregated importance, and current-wave single-domain predictors accounted for 49.66%, compared with 15.66% for interaction terms. Mental Health contributed the largest domain share (26.19%), followed by Friends, Family, and Community (14.60%) and Physical Health (8.40%) (Appendix Table B2). The highest-ranked individual predictors after wave number were weight (5.59%), risk-taking (5.39%), parental knowledge of companions (5.36%), gossiping about another child (4.42%), and friends’ academic achievement (4.38%). Thus, cross-sectional nicotine prediction was concentrated in developmental timing and current-wave behavioral, parenting, peer-context, and physical-health features.

The addition of longitudinal information shifted importance toward temporal features, prior nicotine-use history, and earlier-wave predictors. Together, these longitudinally structured components represented 77.94% of total importance (Appendix Table B2). The top-ranked individual features were wave number (35.18%), nicotine use at the most recent prior wave (8.15%), cumulative prior nicotine-use count (6.34%), and the number of observed prior waves (3.97%). Current-wave single-domain importance declined from 49.66% to 15.06%, and interaction importance declined from 15.66% to 7.00%. Among the retained current-wave interaction terms in the full lagged model, Friends, Family, and Community × Mental Health represented the largest interaction category (2.46% of total beta-aggregated importance), followed by Mental Health × Mental Health (1.81%) and Mental Health × Physical Health (1.74%). Once prior observations were available, nicotine prediction was driven primarily by developmental timing, observation history, and prior nicotine use, replacing the broader current-wave feature profile.

In the lagged substantive analysis, single-domain predictors accounted for 68.27% of retained substantive importance after lagged predictors, temporal variables, and prior-use history variables were excluded. Mental Health represented the largest retained single-domain area (38.50%), followed by Friends, Family, and Community (21.26%) and Physical Health (7.67%). At the individual-feature level, the leading retained features were parental knowledge of companions (7.33%), risk-taking (6.64%), caregiver emotional support (6.38%), feeling interested (5.97%), and weight (4.80%). The largest interaction contributions came from Friends, Family, and Community × Mental Health (11.14%), Mental Health × Mental Health (8.19%), and Mental Health × Physical Health (7.89%) (Appendix Table B2). Thus, although temporal and prior nicotine-use information dominated the full lagged model, the retained current-wave substantive signal was centered on behavioral, emotional, parenting, and peer-context features. Bootstrap stability analyses showed a progressive reduction in the number of stable features across approaches, with 11, six, and three features showing stable positive predictive relevance in the cross-sectional, full lagged, and lagged substantive analyses, respectively (Appendix B, Figures B4–B6).

#### 4.3.3 Marijuana

For marijuana, developmental timing was the most prominent predictive signal in both the cross-sectional and full lagged models, whereas peer-context features dominated the lagged substantive interpretation. Wave number was the dominant individual feature in the cross-sectional model, accounting for 25.34% of total beta-aggregated importance. Current-wave single-domain features accounted for 57.11% of total importance, whereas interaction terms accounted for 17.55%. Among current-wave domains, Mental Health contributed the largest share (23.74%), followed by Friends, Family, and Community (21.92%) and Physical Health (8.23%) (Appendix Table B3). The highest-ranked individual predictors after wave number were having friends who skipped school (11.32%), weight (5.84%), parental knowledge of companions (3.42%), the household-income-by-peer-school-skipping interaction (3.17%), and being the target of peer gossip (2.69%). Thus, cross-sectional marijuana prediction was concentrated in developmental timing together with peer-context, parenting-context, mental-health, and physical-health features.

The addition of longitudinal information increased the prominence of temporal and history-related features, while peer-context information remained visible. In the full lagged model, temporal features accounted for 41.76% of total beta-aggregated importance, prior marijuana-use history accounted for 10.99%, and lagged substantive predictors accounted for 18.83%. Together, these longitudinally structured components represented 71.58% of total importance (Appendix Table B3). The top-ranked individual features were wave number (36.54%), having friends who skipped school (7.77%), the number of prior observed waves (5.22%), cumulative prior marijuana-use count (4.76%), and marijuana use at the most recent prior wave (3.07%). Unlike alcohol and nicotine, where recent prior use ranked immediately behind temporal timing, friends’ school skipping remained the second-highest-ranked marijuana predictor even after longitudinal information was incorporated. Correspondingly, current-wave single-domain importance declined from 57.11% to 21.63%, and current-wave interaction importance declined from 17.55% to 6.82%. Among the retained current-wave interaction terms in the full lagged model, Mental Health × Physical Health represented the largest interaction category (2.29%), followed by Demographics × Mental Health (1.69%) and Demographics × Friends, Family, and Community interactions (1.33%) (Appendix Table B3).

When restricted to current-wave substantive features, single-domain predictors accounted for 76.08% of retained substantive importance. Friends, Family, and Community represented the largest retained single-domain area (36.62%), followed by Mental Health (25.29%) and Physical Health (12.48%). Interaction terms accounted for 23.92% of retained substantive importance, with the largest contributions from Mental Health × Physical Health interactions (8.07%), Demographics × Mental Health interactions (5.93%), and Demographics × Friends, Family, and Community interactions (4.67%) (Appendix Table B3). The top individual retained features were having friends who skipped school (27.35%), weight (7.71%), the temperament/personality item “When I want something, I usually go all the way to get it” (7.06%), being the target of peer gossip (6.24%), and the interaction between that reward-drive item and height (5.77%). Taken together, although the full lagged model was dominated by temporal and prior-use features, the retained current-wave substantive signal was concentrated in peer-context features, with additional contributions from behavioral characteristics, physical-health measures, and interaction terms. Bootstrap stability analyses supported these patterns, with seven, six, and four features showing stable positive predictive relevance in the cross-sectional, full lagged, and lagged substantive analyses, respectively (Appendix B, Figures B7–B9).

## 5. Discussion: Implications, Limitations, and Future Directions

This study contributes a substance-specific longitudinal framework for predicting adolescent alcohol, nicotine, and marijuana use risk across repeated assessments. Incorporating prior-wave information improved prediction, reorganized feature importance, and revealed distinct substance-specific patterns in the current-wave features that remained relevant within history-informed models.

### 5.1.1 Contribution 1: Longitudinal Modeling Improved Prediction and Reorganized Predictive Importance

A primary contribution of this study is the incorporation of lagged predictors and prior assessment history, together with a direct comparison against cross-sectional stacked ensembles. Across all three substances, the lagged stacked ensemble outperformed the corresponding cross-sectional model on both PR-AUC and ROC-AUC. The improvement in PR-AUC was 0.184 for alcohol, 0.183 for nicotine, and 0.144 for marijuana. These gains are especially meaningful given the class imbalance present in each substance-specific outcome.

This contribution extends earlier adolescent substance-use prediction studies. Green et al. (2024) used baseline ABCD information to predict later initiation of any non-prescribed substance. Kim et al. (2025) developed and externally validated a general adolescent substance-use prediction model across three national survey datasets, reporting AUROC values of 0.806, 0.793, and 0.764 across cohorts. The authors identified the lack of longitudinal data and inability to predict future trends as key limitations. More recently, Wei et al. (2026, preprint) compared baseline and dynamic longitudinal multi-task learning models for initiation of alcohol, nicotine, cannabis, and any substance use in ABCD (5.1 release) and reported improved prediction when longitudinal information was incorporated. However, Wei et al. focused on first initiation, whereas the present study modeled subsequent-wave substance-use status across repeated observations. This allowed prior-use history to contribute directly to prediction of both new and repeated positive reports.

Beyond the improvement in discrimination, longitudinal modeling changed the structure of predictive importance. For alcohol, a broad cross-sectional profile characterized by current-wave interactions became less prominent once prior alcohol-use history was included, with cumulative prior use and recent prior use emerging as the leading predictors. For nicotine, developmental timing remained dominant, and the lagged model became strongly concentrated in temporal, prior-use, and prior-wave information. For marijuana, temporal and prior-use information increased in prominence, but peer-context features remained highly ranked. Friends’ school skipping remained the second-highest-ranked predictor in the full lagged model.

### 5.1.2 Contribution 2: Substance-Specific Models Revealed Distinct Predictive Structures

A second contribution is the use of separate predictive models for alcohol, nicotine, and marijuana rather than a single combined substance-use outcome. As noted above, prior work has examined general substance-use initiation (Green et al., 2024) and substance-specific initiation (Wei et al., 2026). The present study addresses a different outcome. It examines subsequent-wave use status separately for each substance under the same cross-sectional and lagged stacked-ensemble procedures. To our knowledge, no prior study has directly compared substance-specific importance structures under both cross-sectional and longitudinal stacked ensemble models for subsequent-wave use status.

The resulting patterns were not interchangeable across substances. For alcohol, prior-use history emerged as the dominant importance area once longitudinal information was included, replacing the broad current-wave interaction profile observed in the cross-sectional model. Nicotine became strongly concentrated in developmental timing and longitudinal structure. Comparatively few current-wave substantive features retained stable positive predictive relevance after history-related variables were excluded from interpretation. Marijuana displayed a different pattern. Although temporal information became prominent, peer-context information remained especially influential, with friends’ school skipping ranking above recent prior marijuana use in the full lagged model and remaining the leading retained substantive feature.

These distinctions matter because a combined substance-use outcome may conceal differences in the predictive organization of risk. Prior literature has identified previous drinking, peer context, parenting factors, and behavioral characteristics as relevant to adolescent substance use (Mason et al., 2017; Mantey et al., 2022; Stautz & Cooper, 2013). The present study adds evidence that their relative importance differs across alcohol, nicotine, and marijuana once repeated assessment history is incorporated. This supports the use of separate prediction models when the objective is to identify the information most relevant to each substance’s risk profile.

### 5.1.3 Contribution 3: Lagged Substantive Interpretation Revealed How Current-Wave Features Changed Within History-Informed Models

A third contribution is the identification of current-wave substantive features that remained prominent after longitudinal history was incorporated. In the full lagged models, prior-use history and temporal variables were expected to become powerful predictors. However, these variables provide limited information about the contemporaneous social, behavioral, and health-related characteristics associated with elevated predicted risk. The lagged substantive analysis addressed this problem by filtering history-related variables from the importance ranking while retaining the fitted longitudinal model. This analysis did not estimate a separate reduced model. Instead, it identified the current-wave signals whose relevance was evaluated within a model already informed by prior history. This interpretive approach has not been applied in prior adolescent substance-use prediction studies, which have generally reported importance rankings without distinguishing longitudinal history variables from current-wave substantive signals.

For alcohol, the retained current-wave profile changed from the broad peer/family and behavioral interaction pattern observed in the cross-sectional model toward a profile characterized by physical-development and contextual interaction features. Weight, height-by-weight, peer-shoplifting-by-height, and height-by-parental-knowledge were among the most prominent retained features, while positive urgency showed stable positive predictive relevance. Prior meta-analytic evidence has associated positive urgency and sensation seeking with adolescent alcohol use (Amialchuk et al., 2021; Stautz & Cooper, 2013). The present findings extend this literature by showing that positive urgency and selected contextual interaction signals remained informative within an alcohol prediction model that incorporated prior alcohol-use history.

For nicotine, the retained current-wave profile became substantially narrower after longitudinal information was incorporated. The cross-sectional model included behavioral, parenting, peer-context, and physical-health features, whereas the lagged substantive analysis identified only three retained features with stable positive predictive relevance: caregiver emotional support, gossiping about another child, and having friends who were excellent students. Mantey et al. (2022) reported that higher perceived parental knowledge was associated with lower odds of nicotine-vaping initiation. Doran et al. (2025) examined social-cognitive influences on susceptibility to nicotine and tobacco use over time in ABCD. The present study adds that, within the history-informed nicotine model, the stable current-wave substantive profile was narrow, concentrated in selected parenting and peer-context indicators.

For marijuana, peer-context information remained prominent despite the incorporation of longitudinal history. In both the cross-sectional and lagged substantive interpretations, friends’ school skipping was a leading feature, and it remained the second-highest-ranked feature in the full lagged model. Peer-gossip victimization also remained a stable retained substantive signal. Prior longitudinal research has demonstrated that close friends’ substance use, offers to use substances, and risky behaviors predict later adolescent cannabis use (Mason et al., 2017). The present study extends this evidence by showing that peer school disengagement remained highly prominent within the history-informed marijuana model. Taken together, these findings reveal that the longitudinal models captured substance-specific current-wave patterns beyond temporal and prior-use signals.

### 5.1.4 Contribution 4: Participant-Level Stability Analysis Distinguished High-Ranking Features From Features With Stable Positive Predictive Relevance

A fourth contribution is the evaluation of feature-importance stability through participant-level bootstrap analysis. Prior machine-learning studies of adolescent substance use have reported predictive performance and feature-importance results, including SHAP-based identification of influential variables (Kim et al., 2025). To our knowledge, participant-level bootstrap stability analysis has not previously been applied to evaluate feature-importance reproducibility in adolescent substance-use prediction models. The present study adds a complementary stability evaluation by examining whether permuting each top-ranked feature consistently reduced the PR-AUC of the complete stacked ensemble across participant-level resamples. This distinction is important because a feature may rank highly in a fitted importance analysis without showing stable positive predictive relevance across resampled adolescents.

The stability results differed across both modeling approaches and substances. Alcohol showed the broadest stable feature profile across all three analyses, with 14, 13, and 11 of the top 20 features showing stable positive predictive relevance in the cross-sectional, full lagged, and lagged substantive analyses, respectively. Nicotine showed the greatest reduction in the number of stable top-ranked features across approaches, decreasing from 11 stable features in the cross-sectional analysis to six in the full lagged analysis and three in the lagged substantive analysis. Marijuana showed seven stable features in the cross-sectional analysis, six in the full lagged analysis, and four in the lagged substantive analysis. These findings indicate that, although longitudinal modeling improved predictive performance across all three substances, the retained current-wave substantive interpretation became more selective, particularly for nicotine. These patterns reinforce the substance-specific findings described in Contribution 3, with nicotine showing the greatest narrowing of stable current-wave relevance after longitudinal information was incorporated.

Consistent with broader principles for interpretable machine learning, model explanations should be evaluated in relation to their scientific purpose and should not be treated as definitive evidence solely because a feature ranks highly in one fitted model (Murdoch et al., 2019). Features that were both highly ranked and showed stable positive predictive relevance therefore warrant greater interpretive emphasis and future validation than features that ranked highly but had stability intervals including zero. The present study adds a participant-level stability evaluation of top-ranked features within complete stacked ensembles, providing a stronger basis for identifying predictors whose relevance is reproducible across participant-level resamples. This distinction is particularly relevant for prevention-oriented risk screening, where predictive features may help guide further assessment or outreach but should not be treated as causal or directly actionable targets.

### 5.1.5 Overall Contribution

Overall, this study extends adolescent substance-use prediction research by demonstrating that predictive importance is both substance-specific and dependent on the availability of longitudinal history. Across substances, longitudinal information reorganized predictive importance in distinct ways, with alcohol becoming history-dominated, nicotine concentrating in temporal structure, and marijuana retaining a distinctive peer-context signal. These findings are predictive rather than causal, but they provide a foundation for future validation of substance-specific risk-targeting approaches for adolescent prevention.

#### 5.2 Implementation Considerations and Limitations

Although these models show promise for prevention-oriented adolescent substance-use risk screening, model outputs should be interpreted with expert oversight and used to support outreach or preventive services rather than to stigmatize or label adolescents. Seven limitations should be noted when interpreting the findings.

### 5.2.1 Sample Generalizability and External Validation

The generalizability of the findings is limited by two aspects of the ABCD sampling design. The ABCD Study recruited participants through 21 research sites distributed across the United States and was designed to approximate national sociodemographic diversity (Garavan et al., 2018). However, it was not designed to represent every geographic, cultural, or environmental context equally. The ABCD sample also does not perfectly reproduce the racial, ethnic, socioeconomic, and caregiver-education distributions of the broader U.S. adolescent population (Barch et al., 2018). Substance-use patterns and predictive relationships in regions or populations less well represented in the cohort may therefore differ from those captured in the present models. This issue is particularly important for interpreting features involving demographic characteristics or contextual interactions, which should not be treated as inherent individual-level risk mechanisms.

Additionally, the models were not externally validated in an independent adolescent cohort, and such validation is needed before the models or their identified predictive patterns are considered for prevention-oriented implementation. Participant-level train-test separation and bootstrap stability analyses assessed predictive performance and feature-level stability within the present sample. However, these procedures do not establish transportability to other adolescent populations, geographic settings, health systems or school systems. Future work should evaluate model performance and feature-importance patterns in geographically and demographically diverse adolescent samples.

### 5.2.2 Predictor Availability

Real-world applicability may be limited by predictor availability. The models relied on a comprehensive set of predictors collected across repeated ABCD assessments, including behavioral, peer, family, mental-health, physical-health, and substance-history information. Many of these measures may not be routinely available in clinical, school, or public-health settings, particularly in systems without access to detailed longitudinal assessment records (Rakers et al., 2024; Watson et al., 2020). Practical deployment would therefore require evaluation of reduced predictor sets feasible to collect in routine screening workflows. These sets would need to preserve adequate predictive performance.

### 5.2.3 Developmental Stage

The developmental stage of participants may have affected the predictive contribution of some mental-health features. Participants entered the ABCD Study in late childhood and were followed through adolescence, a developmental period during which psychiatric symptoms and behavioral vulnerabilities may emerge or change substantially. Because some symptoms may not be fully expressed at earlier assessments, their predictive contribution to subsequent-wave substance-use status may be underestimated within the observed study window (Kessler et al., 2007). Future analyses using later ABCD waves may determine whether mental-health features become more prominent as the cohort progresses through adolescence.

### 5.2.4 Outcome Definition

A limitation of the present outcome definition is that it did not distinguish first initiation from repeated substance-use reports at later assessment waves. The models predicted whether an adolescent reported substance use at a subsequent wave. A positive outcome could therefore represent either a first reported use or a repeated positive report among adolescents with prior use history. This definition did not isolate predictors associated specifically with continued use, escalation, or the development of substance-use disorder. Future work should examine these outcomes separately because they represent distinct prevention and intervention questions. This distinction is also important when comparing the present study with recent longitudinal ABCD research focused specifically on first initiation (Wei et al., 2026).

### 5.2.5 Missing Data

Missing-data handling may introduce uncertainty into the findings. Training-only imputation procedures were applied to reduce information leakage. However, imputed values may still influence predictive patterns when missingness is systematic or related to unmeasured participant characteristics. The results should therefore be interpreted as patterns estimated from a combination of observed and imputed predictor values. Future sensitivity analyses should evaluate whether the major predictive and feature-importance findings remain consistent under alternative missing-data approaches.

### 5.2.6 Feature-Importance Interpretation

The feature-importance and stability findings should be interpreted as exploratory predictive evidence. Permutation-based importance may be influenced by correlation among predictors because related features can share or substitute for predictive information. In addition, the 95% stability intervals characterize stability among selected top-ranked features within the present data but do not provide confirmatory inference for all predictors or support generalization beyond the present sample. The lagged substantive analyses filtered the interpretation ranking from the fitted full lagged models rather than estimating new reduced models. Consequently, retained substantive features and interaction patterns should be prioritized for sensitivity analysis and replication before being considered for prevention or intervention planning.

#### 5.3 Future Directions

First, future work could adapt the stacking framework for initiation prediction by redefining the outcome as first observed use and constructing lagged features that capture pre-initiation risk trajectories (Wei et al., 2026). This would target a different and arguably more prevention-relevant stage of adolescent substance-use development (Matson et al., 2022).

Second, the feature-tiering thresholds used in Phase 2 were derived from the alcohol importance distribution, as alcohol had the most balanced class ratio among the three substances. This carries an implicit assumption that a single set of thresholds is appropriate across all substances. Feature importance estimates are sensitive to class imbalance, with more balanced distributions producing more stable and reliable rankings (Dube & Verster, 2023; Tharwat, 2021). In practice, nicotine and marijuana have substantially lower positive-class prevalence. Their importance distributions may therefore be compressed toward zero, making the alcohol-derived thresholds potentially overly conservative. Future work could explore substance-specific threshold calibration, allowing each substance to define its own tiering boundaries and potentially retaining more informative features for rarer outcomes.

Third, the current pipeline used XGBoost as the sole screening model for feature importance ranking. The retained feature set is therefore sensitive to this choice, as different models may identify different predictors as informative. Future work could compare alternative screening approaches, such as elastic net logistic regression, random forest-based importance, or LASSO-based screening. This would assess whether the retained feature sets are robust across methods.

Fourth, the stacking architecture could be expanded to include recurrent neural networks or long short-term memory models. These architectures are specifically designed for sequential data and may better capture temporal patterns across waves than the current base learners. Incorporating these models as additional base learners would allow the meta-learner to combine tree-based, linear, neural, and sequential predictive signals. This could improve both performance and interpretability across substances.

## Data Availability

Data used in the preparation of this article were obtained from the Adolescent Brain Cognitive Development (ABCD) Study (https://abcdstudy.org). The ABCD data repository grows and changes over time. The ABCD data used in this report came from Release [6.0], https://nda.nih.gov/abcd.

## Appendix A

### Model Hyperparameter Specifications

All models were implemented using Python. Hyperparameter settings were manually specified for training-only feature screening rather than final predictive optimization and were held constant across all three substances (alcohol, nicotine, and marijuana).

**Table A1.**
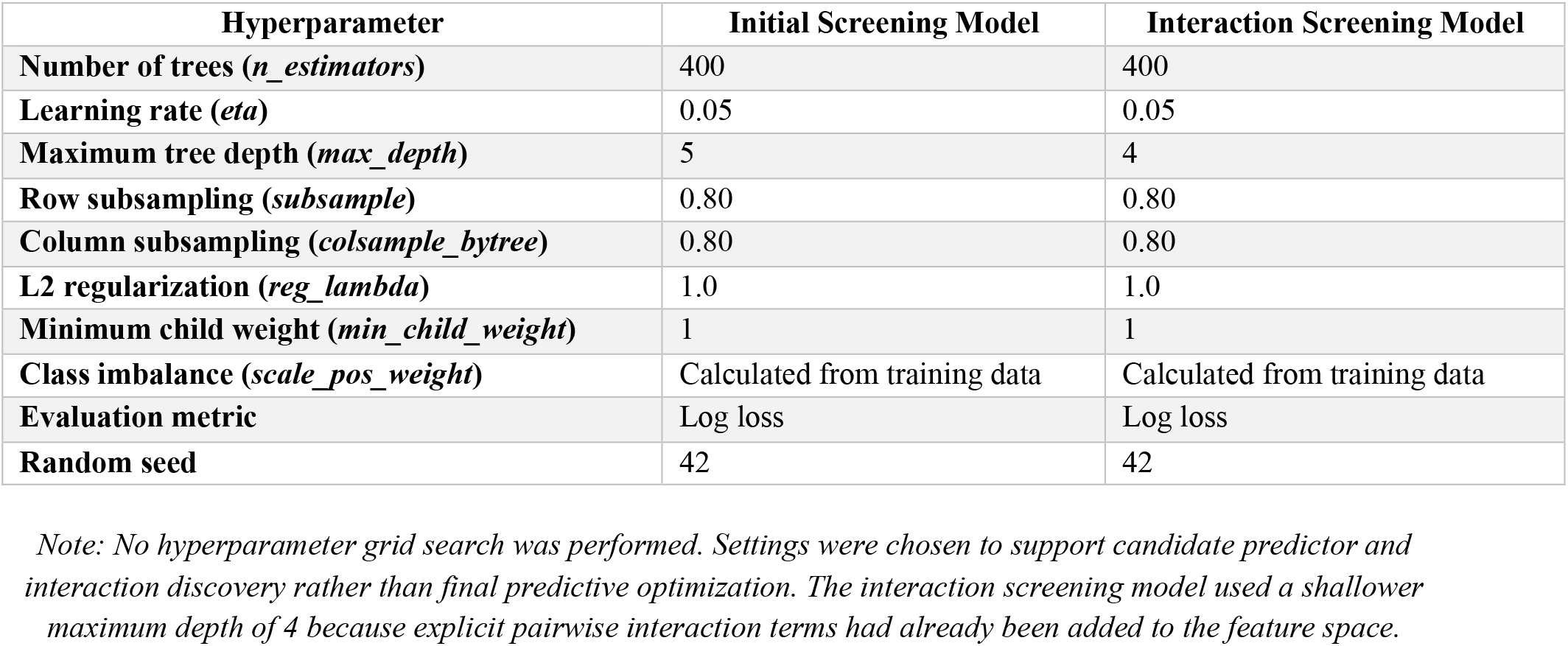
XGBoost Hyperparameter Specifications for Screening Models.

| Hyperparameter | Initial Screening Model | Interaction Screening Model |
| --- | --- | --- |
| Number of trees (*n_estimators*) | 400 | 400 |
| Learning rate (*eta*) | 0.05 | 0.05 |
| Maximum tree depth (*max_depth*) | 5 | 4 |
| Row subsampling (*subsample*) | 0.80 | 0.80 |
| Column subsampling (*colsample_bytree*) | 0.80 | 0.80 |
| L2 regularization (*reg_lambda*) | 1.0 | 1.0 |
| Minimum child weight (*min_child_weight*) | 1 | 1 |
| Class imbalance (*scale_pos_weight*) | Calculated from training data | Calculated from training data |
| Evaluation metric | Log loss | Log loss |
| Random seed | 42 | 42 |

*Note: No hyperparameter grid search was performed. Settings were chosen to support candidate predictor and interaction discovery rather than final predictive optimization. The interaction screening model used a shallower maximum depth of 4 because explicit pairwise interaction terms had already been added to the feature space.*

**Table A2.**
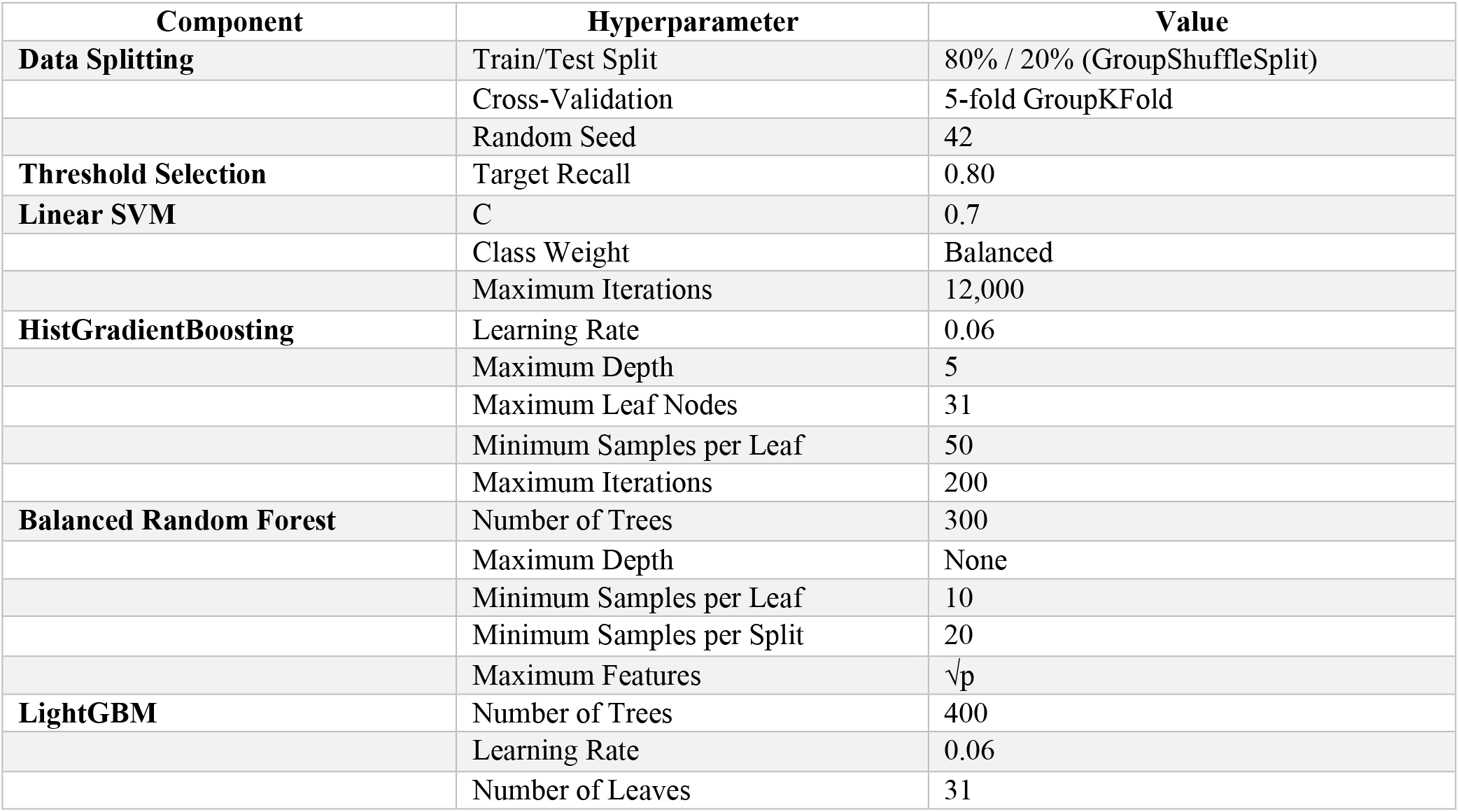
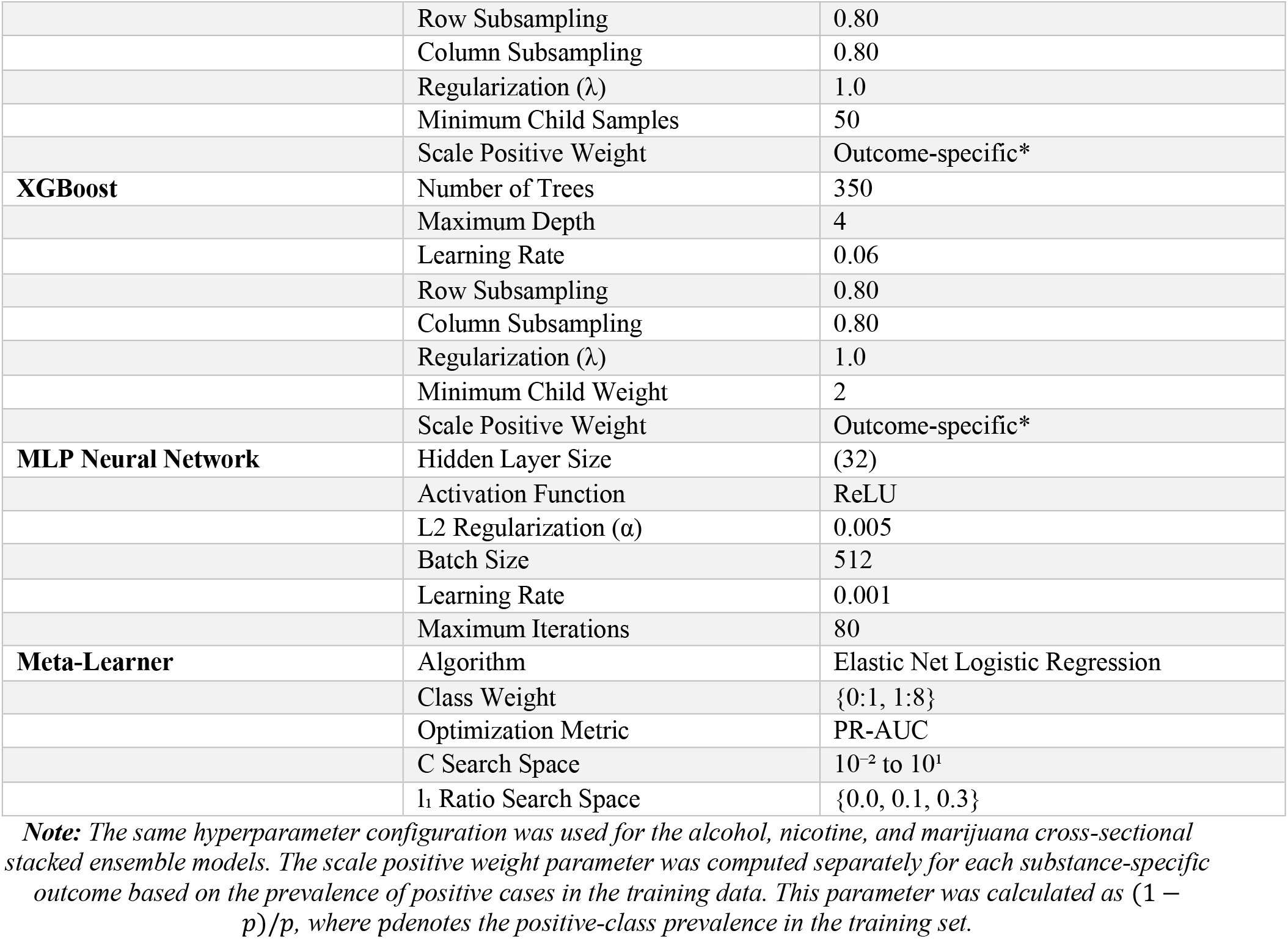
Hyperparameter Specifications for Cross-sectional and Lagged Stacked Ensembles.

**Table A3.** Additional Longitudinal Hyperparameter Specifications for the Lagged Stacked Ensemble Models.

| Component | Specification |
| --- | --- |
| **Maximum lag depth** | 4 waves |
| **Lagged predictors** | Lag1–Lag4 for all selected raw and interaction-source features |
| **Prior-use indicators** | Previous-use status at the prior four waves |
| **Cumulative count feature** | Number of prior substance-use occurrences |
| **Cumulative proportion feature** | Proportion of prior waves with substance use |
| **Ever-use indicator** | Whether substance use occurred in any previous wave |
| **Prior wave count** | Number of available prior observations |
| **Temporal feature** | Wave number (*wave_num*) |
| **Minimum history requirement** | At least one prior wave available (Lag1) |
| **Interaction reconstruction** | Top 20 interaction terms rebuilt after preprocessing and encoding |

*Note: The lagged stacked ensemble models used the same machine-learning algorithms, hyperparameter settings, cross-validation strategy, and threshold-selection procedure as the cross-sectional stacked ensemble models (Table A2). Additional longitudinal information was incorporated through lagged predictors, substance-specific prior-use history variables, cumulative exposure measures, and temporal features. Lagged features were constructed from the previous four waves, and observations were required to have at least one prior wave available for inclusion in the analysis.*

## Appendix B

**Table B1.** Feature Domain Contributions Across Alcohol-Use Prediction Models.

| Feature Area | Cross-Sectional Model | Full Lagged Model | Lagged Substantive Analysis |
| --- | --- | --- | --- |
| Prior alcohol-use history | — | 48.93% | — |
| Temporal / developmental timing | 8.59% | 11.06% | — |
| Temporal / observation history | — | 1.60% | — |
| *Current-wave single-domain features* |  |  |  |
| Demographics | 3.83% | 0.20% | 0.73% |
| Friends, Family & Community | 12.12% | 1.93% | 7.07% |
| Mental Health | 17.87% | 3.87% | 14.19% |
| Physical Health | 6.05% | 4.14% | 15.18% |
| Study site / design variable | 1.90% | 0.99% | 3.62% |
| *Current-wave interaction features* |  |  |  |
| Demographics × Mental Health | 2.29% | 0.41% | 1.50% |
| Demographics × Physical Health | 2.02% | 2.56% | 9.38% |
| Friends, Family & Community × Mental Health | 15.76% | 2.17% | 7.98% |
| Friends, Family & Community × Physical Health | 8.21% | 4.27% | 15.67% |
| Friends, Family & Community × Study Site / Design Variable | 1.54% | 0.36% | 1.34% |
| Mental Health × Physical Health | 10.25% | 3.24% | 11.89% |
| Physical Health × Physical Health | 9.58% | 3.12% | 11.45% |
| *Lagged substantive predictors* |  |  |  |
| Lagged: Demographics | — | 0.89% | — |
| Lagged: Friends, Family & Community | — | 2.81% | — |
| Lagged: Mental Health | — | 4.51% | — |
| Lagged: Physical Health | — | 1.99% | — |
| Lagged: Study Site / Design Variable | — | 0.96% | — |
| **Total** | **100.00%** | **100.00%** | **100.00%** |

*Note.* Percentages reflect each feature area's share of total predictor contribution within model. Em dashes (—) indicate the feature area is not applicable to that model specification.

**Figure B1.**
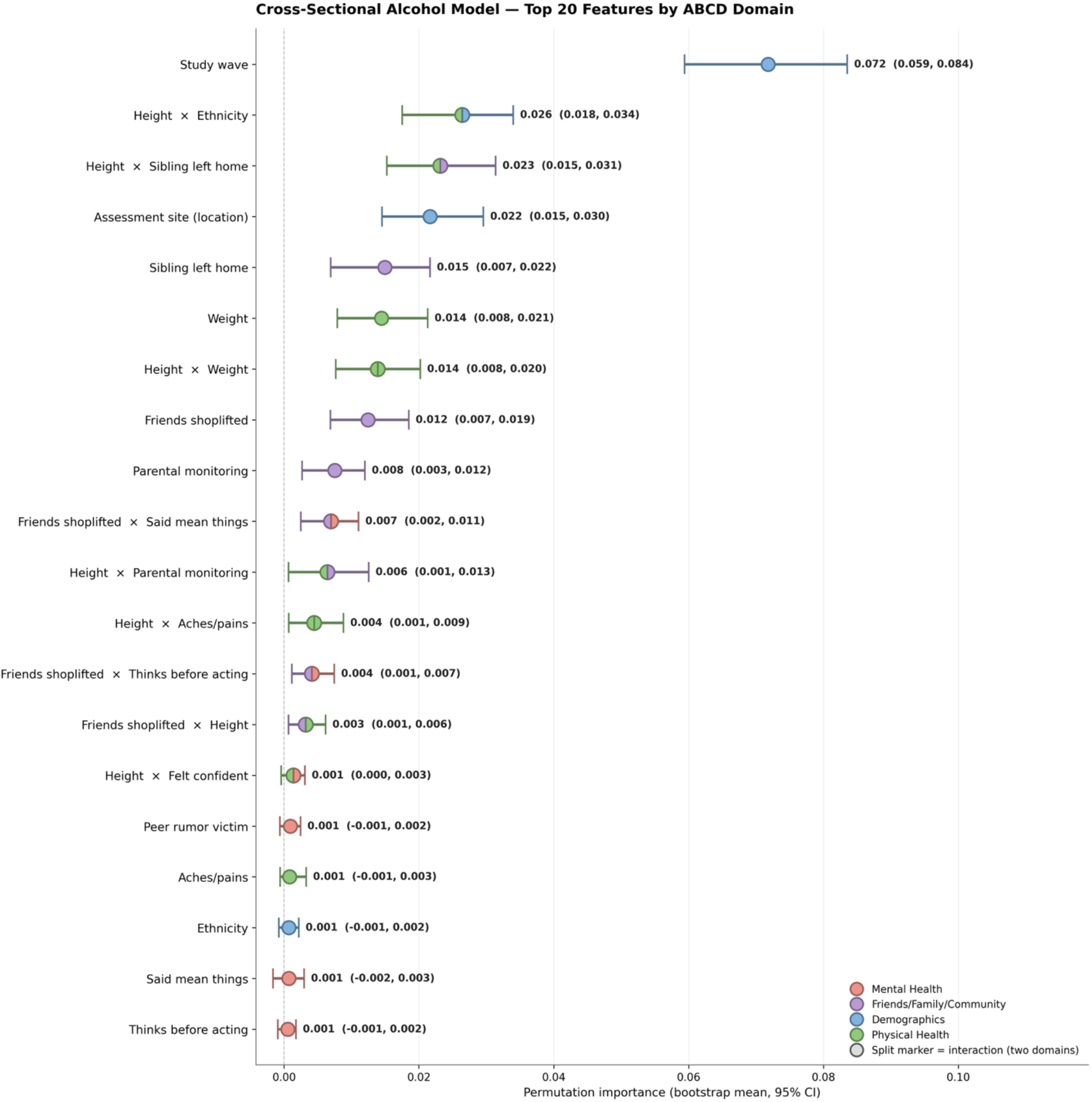
Alcohol Cross-Sectional Analysis: Stacked-Model Permutation Importance and 95% Stability Intervals for the Top 20 Features.

**Figure B2.**
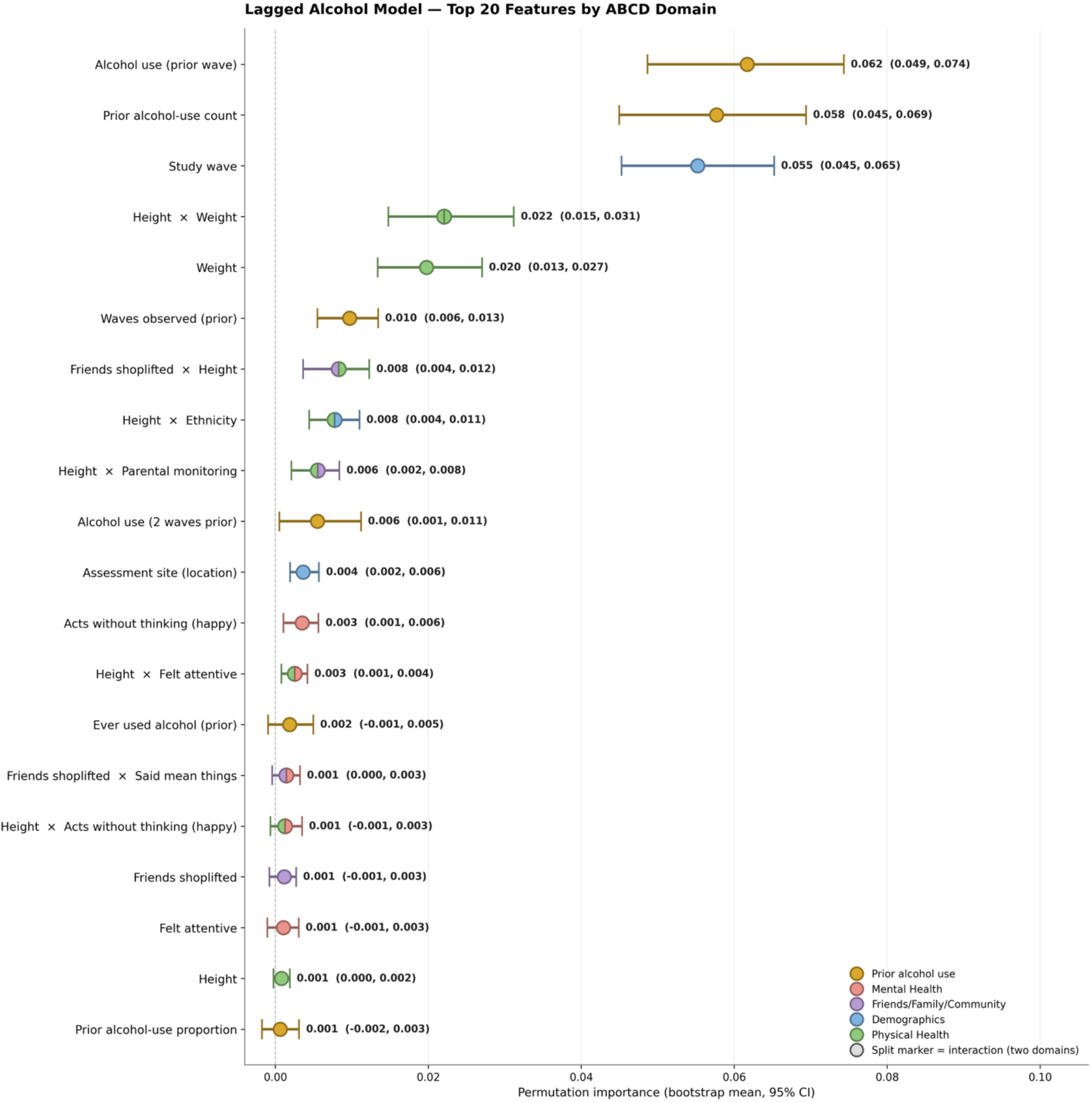
Alcohol Full Lagged Analysis: Stacked-Model Permutation Importance and 95% Stability Intervals for the Top 20 Features.

**Figure B3.**
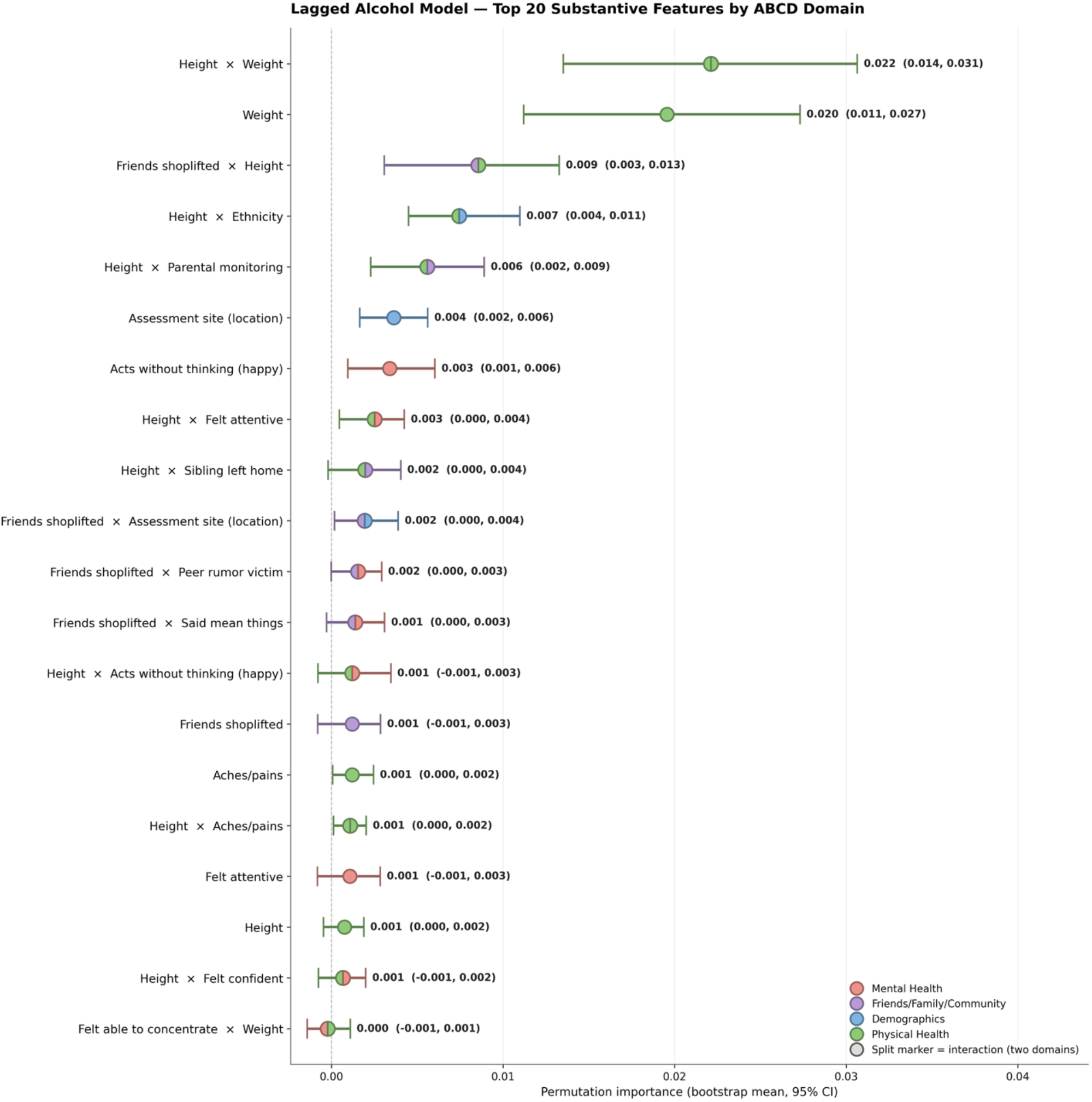
Alcohol Lagged Substantive Analysis: Stacked-Model Permutation Importance and 95% Stability Intervals for the Top 20 Retained Features.

**Table B2.**
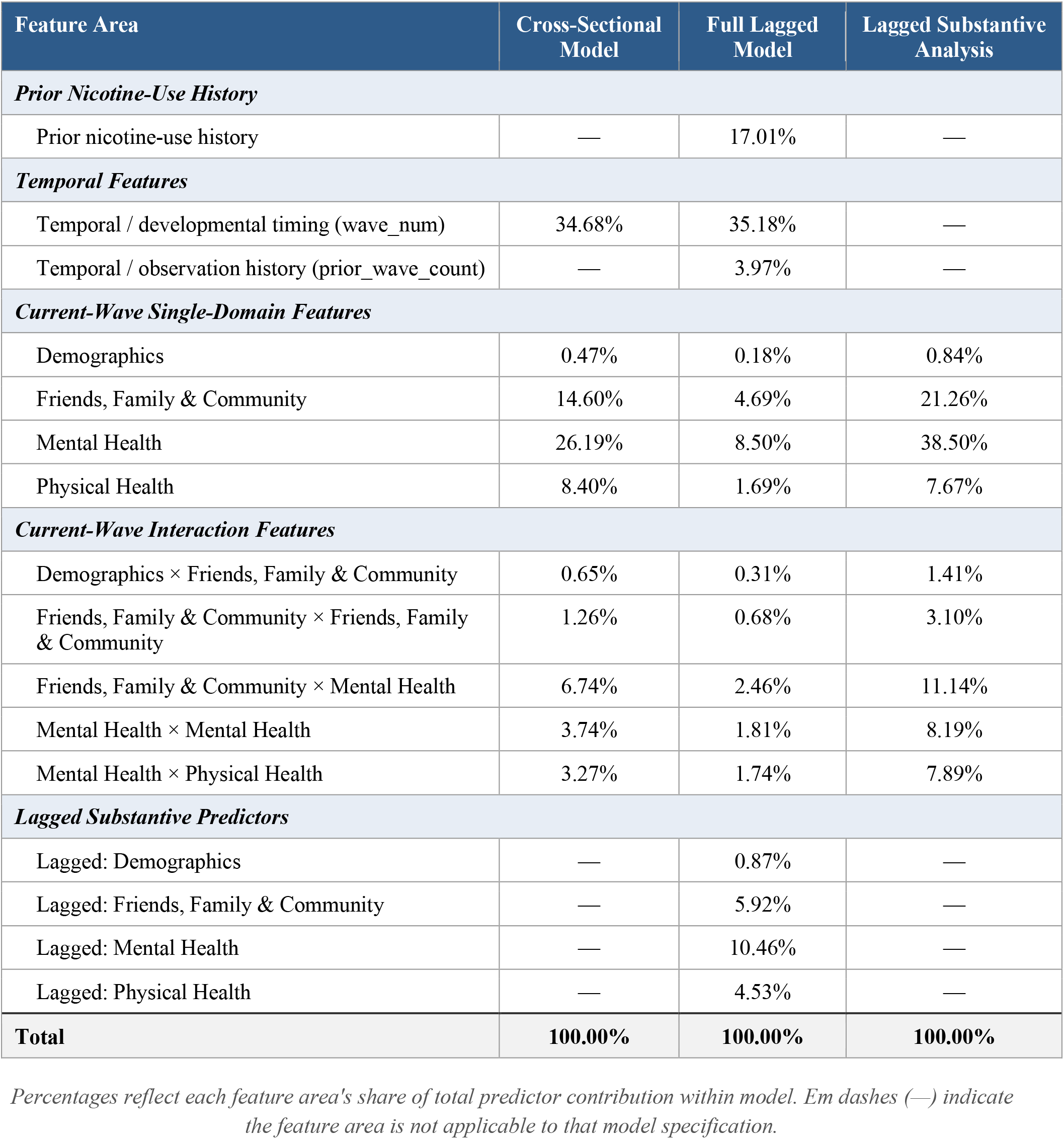
Beta-Aggregated Feature Importance Contributions Across Nicotine-Use Prediction Models.

**Figure B4.**
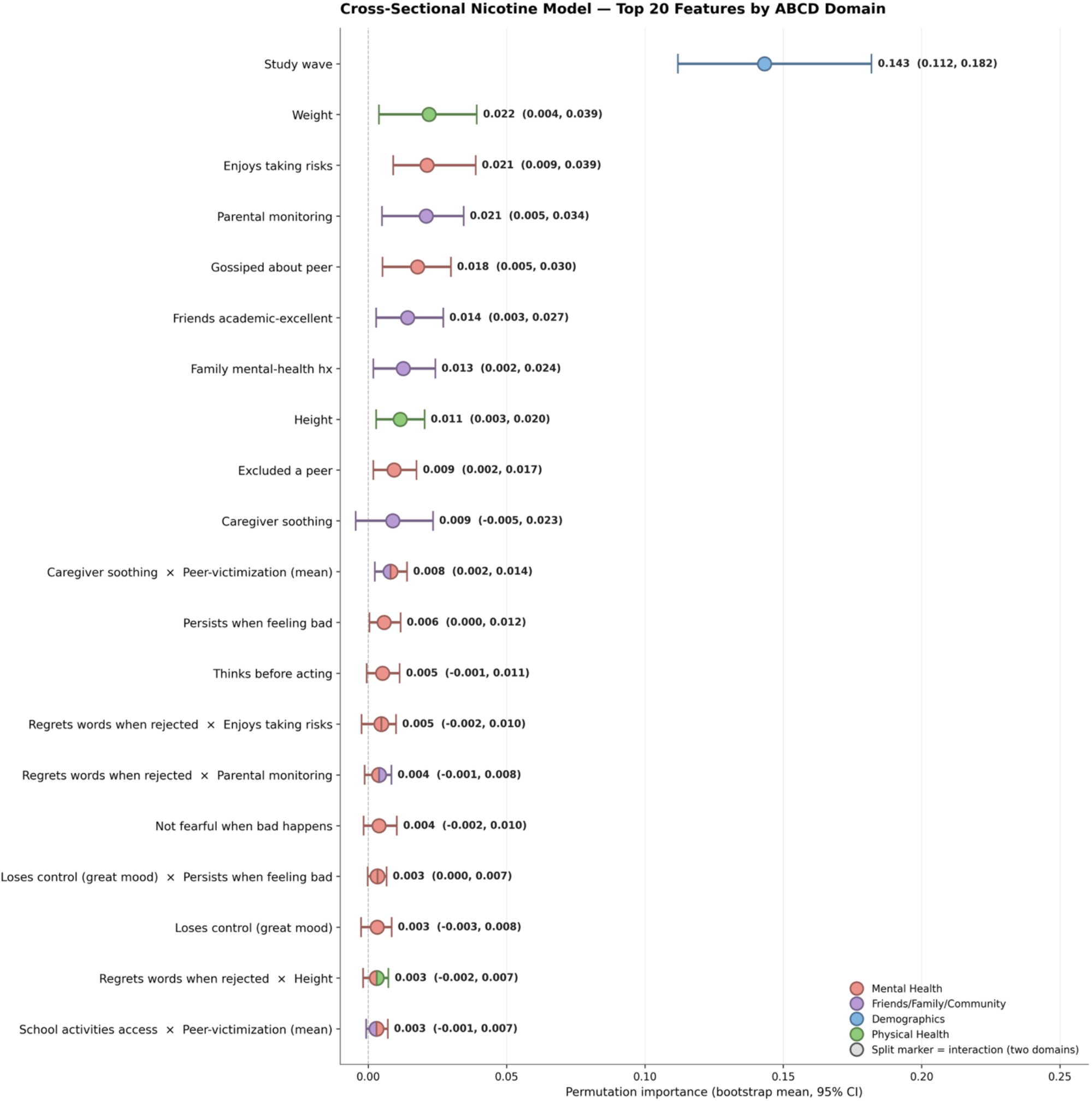
Nicotine Cross-Sectional Analysis: Stacked-Model Permutation Importance and 95% Stability Intervals for the Top 20 Features.

**Figure B5.**
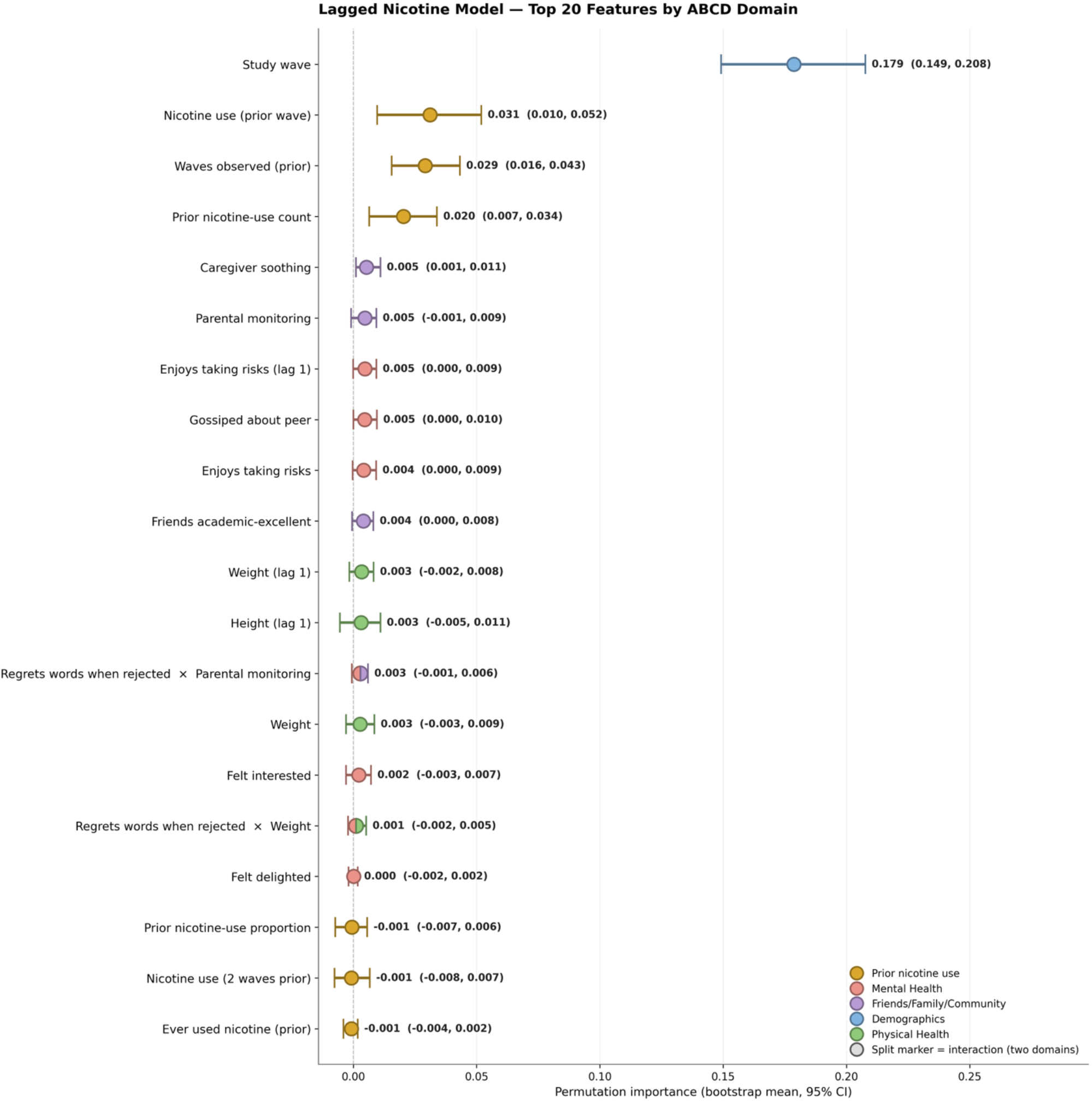
Nicotine Full Lagged Analysis: Stacked-Model Permutation Importance and 95% Stability Intervals for the Top 20 Features.

**Figure B6.**
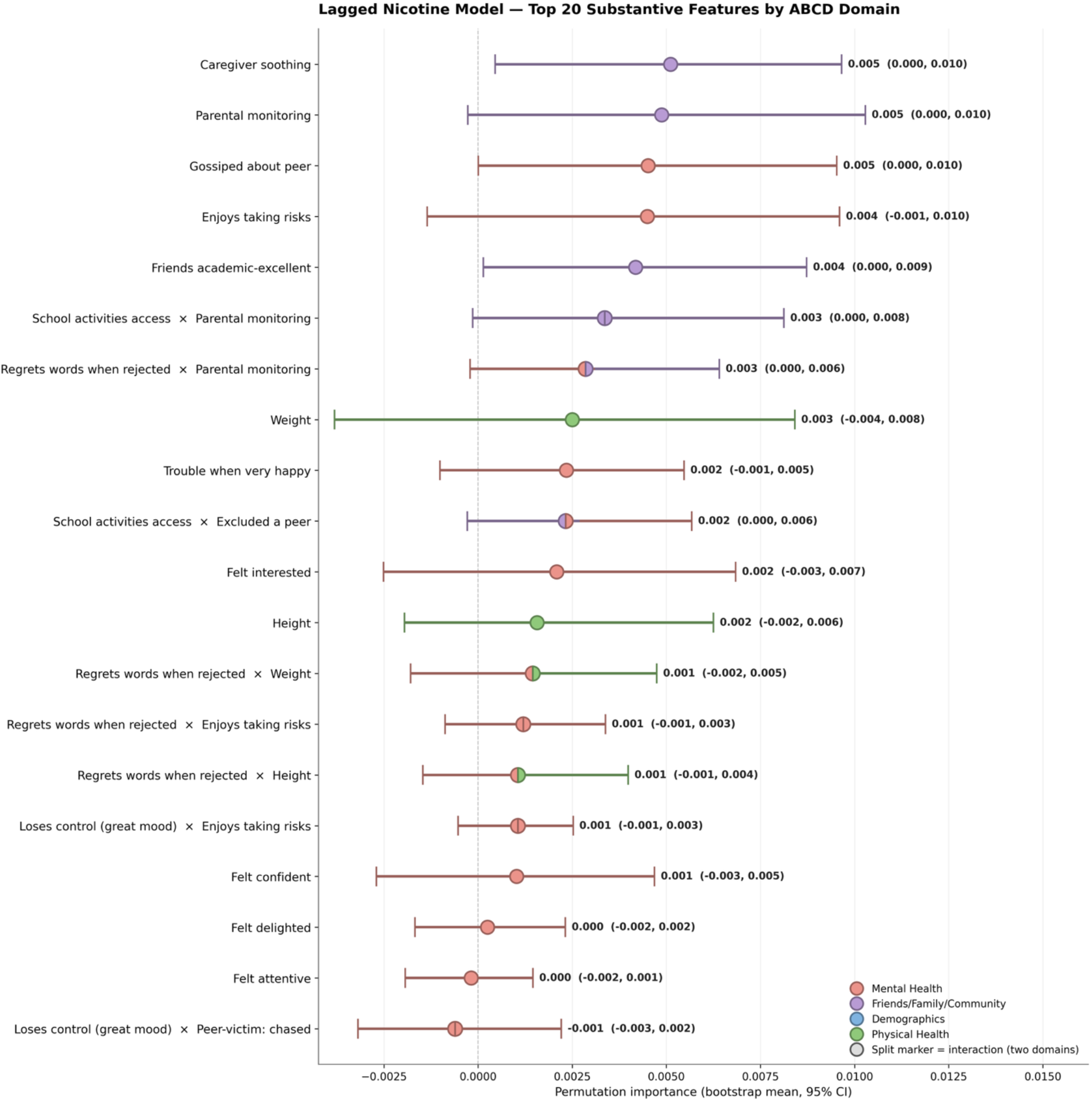
Nicotine Lagged Substantive Analysis: Stacked-Model Permutation Importance and 95% Stability Intervals for the Top 20 Retained Features.

**Appendix Table B3.** Beta-Aggregated Feature Importance by Feature Area Across Marijuana Modeling Approaches.

| Feature Area | Cross-Sectional Model | Full Lagged Model | Lagged Substantive Analysis |
| --- | --- | --- | --- |
| ***Prior Marijuana-Use History*** |  |  |  |
| Prior marijuana-use history | — | 10.99% | — |
| ***Temporal Features*** |  |  |  |
| Temporal / developmental timing (wave_num) | 25.34% | 36.54% | — |
| Temporal / observation history (prior_wave_count) | — | 5.22% | — |
| ***Current-Wave Single-Domain Features*** |  |  |  |
| Demographics | 3.22% | 0.48% | 1.69% |
| Friends, Family & Community | 21.92% | 10.41% | 36.62% |
| Mental Health | 23.74% | 7.19% | 25.29% |
| Physical Health | 8.23% | 3.55% | 12.48% |
| ***Current-Wave Interaction Features*** |  |  |  |
| Demographics × Friends, Family & Community | 3.17% | 1.33% | 4.67% |
| Demographics × Mental Health | 7.31% | 1.69% | 5.93% |
| Demographics × Physical Health | 2.19% | 0.60% | 2.10% |
| Friends, Family & Community × Mental Health | 0.79% | 0.46% | 1.61% |
| Mental Health × Mental Health | 1.87% | 0.44% | 1.53% |
| Mental Health × Physical Health | 2.23% | 2.29% | 8.07% |
| ***Lagged Substantive Predictors*** |  |  |  |
| Lagged: Demographics | — | 2.80% | — |
| Lagged: Friends, Family & Community | — | 5.39% | — |
| Lagged: Mental Health | — | 7.06% | — |
| Lagged: Physical Health | — | 3.58% | — |
| **Total** | **100.00%** | **100.00%** | **100.00%** |

*Percentages reflect each feature area's share of total predictor contribution within model. Em dashes (—) indicate the feature area is not applicable to that model specification.*

**Figure B7.**
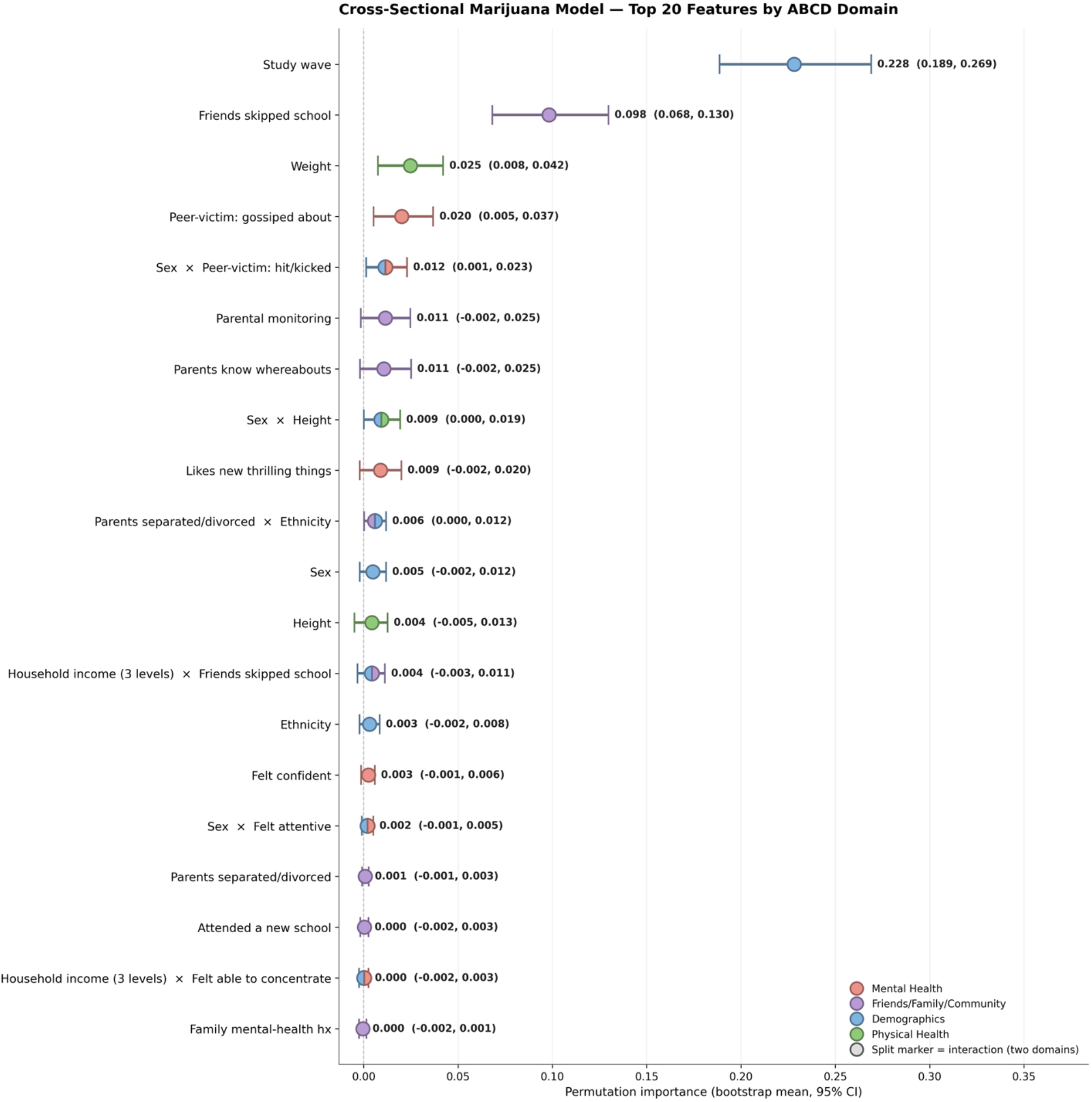
Marijuana Cross-Sectional Analysis: Stacked-Model Permutation Importance and 95% Stability Intervals for the Top 20 Features.

**Figure B8.**
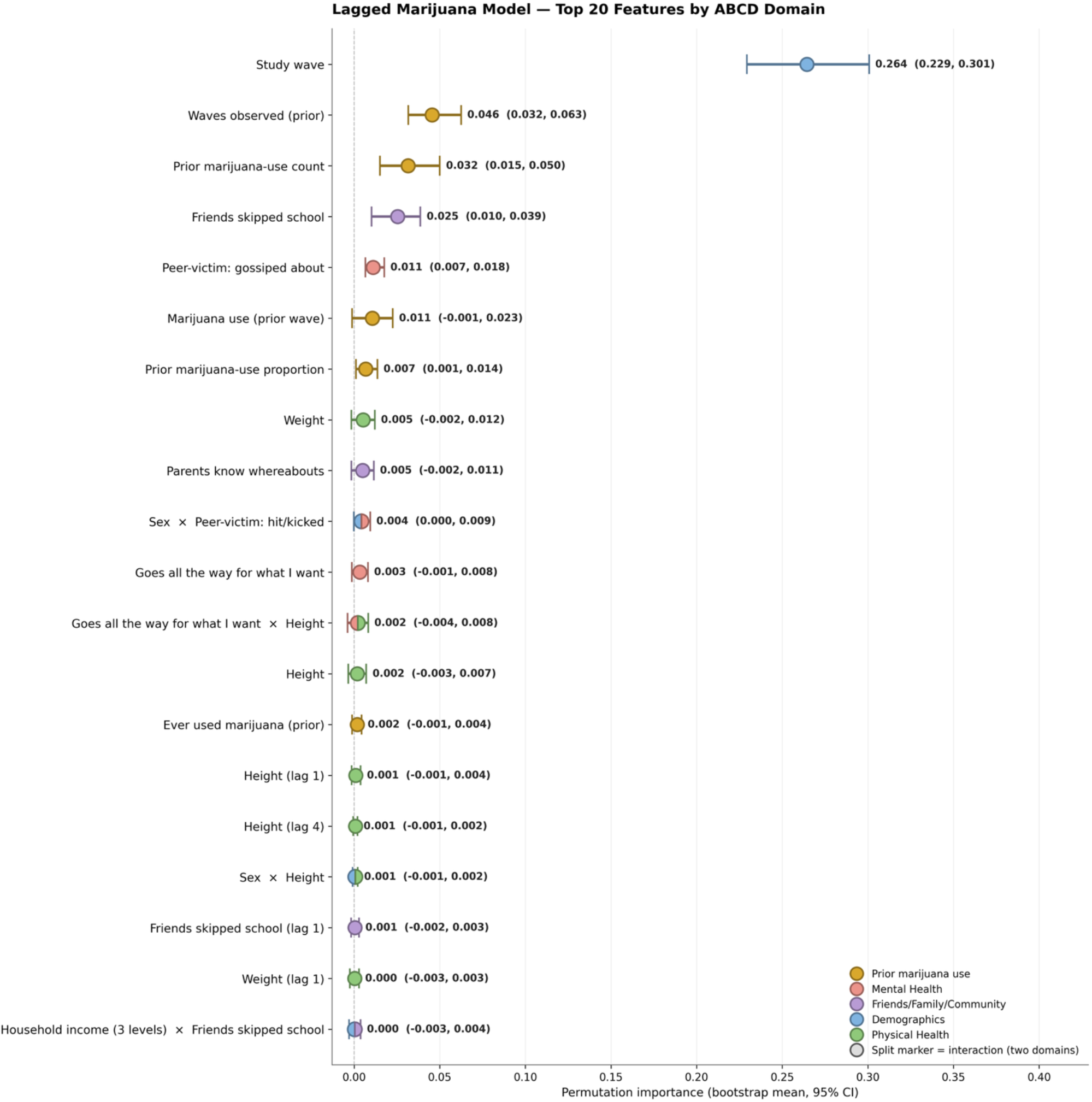
Marijuana Full Lagged Analysis: Stacked-Model Permutation Importance and 95% Stability Intervals for the Top 20 Features.

**Figure B9.**
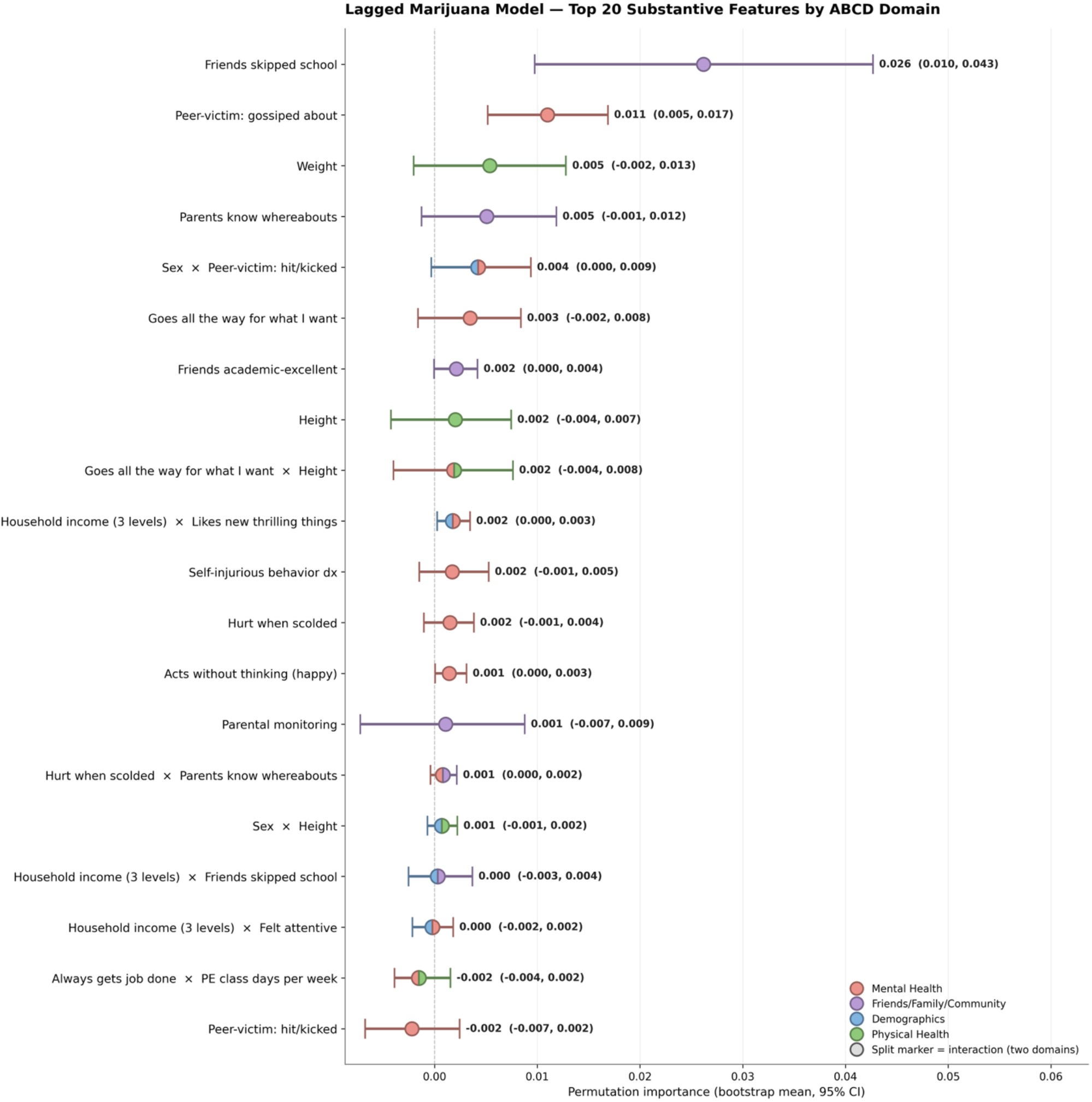
Marijuana Lagged Substantive Analysis: Stacked-Model Permutation Importance and 95% Stability Intervals for the Top 20 Retained Features.

## Appendix C

**Table C1.** Distribution of predictor variables across conceptual domains.

| Domain | Subcategory | n | % of total |
| --- | --- | --- | --- |
| Demographics | Parent's occupation | 2 | 0.6% |
|  | General child demographics | 12 | 3.7% |
|  | *Domain subtotal* | **14** | **4.3%** |
| Friends, Family & Community | Culture | 2 | 0.6% |
|  | Family environment | 9 | 2.8% |
|  | Parenting behavior | 16 | 5.0% |
|  | Peers | 6 | 1.9% |
|  | Temperament / personality | 3 | 0.9% |
|  | Work and school | 12 | 3.7% |
|  | *Domain subtotal* | **48** | **14.9%** |
| Physical Health | Anthropometrics and vitals | 4 | 1.2% |
|  | Medical / pain variables | 1 | 0.3% |
|  | Physical activity | 3 | 0.9% |
|  | *Domain subtotal* | **8** | **2.5%** |
| Mental Health | Anxiety | 38 | 11.8% |
|  | Emotion / Positive Effect | 9 | 2.8% |
|  | Harm / suicidality | 47 | 14.6% |
|  | Mood / depressive disorder | 49 | 15.2% |
|  | Peer behavior | 19 | 5.9% |
|  | Psychotic behavior | 21 | 6.5% |
|  | Sleep problems | 4 | 1.2% |
|  | Stress levels | 25 | 7.7% |
|  | Temperament / personality | 41 | 12.6% |
|  | *Domain subtotal* | **253** | **78.3%** |
| **Total predictor variables** |  | **323** | **100.0%** |

*Note. Predictors derived from the ABCD Study (v6.0) are organized into four conceptual domains and 21 subcategories. Percentages reflect each subcategory's share of the total predictor pool (N = 325). Domain subtotals are shown in bold italics. The "temperament/personality" subcategory appears in two domains (Friends, Family & Community; Mental Health) because items differ by source and construct.*

**Table C2.** Predictor Variable Inventory. *Complete list of predictors organized by domain and subcategory (ABCD Study v6.0)*

| **Domain: Demographics (n = 14)** |  |
| --- | --- |
| **#** | **Variable** |
| **Subcategory: Parent's Occupation (n = 2)** |  |
| 1 | parent's occupation industry |
| 2 | parent's occupation classification |
| **Subcategory: General Child Demographics (n = 12)** |  |
| 1 | CohortDate of birth (Anonymized) |
| 2 | Age |
| 3 | Ethnicity |
| 4 | Hispanic |
| 5 | Sex |
| 6 | Current school grade |
| 7 | Caregivers Highest education |
| 8 | Caregivers' partnership and employment status |
| 9 | Household income - 6 levels |
| 10 | Household income - 3 levels |
| 11 | Informant's relationship to child |
| 12 | State |

Friends, Family & Community
| Domain: Friends, Family & Community (n = 48) |  |
| --- | --- |
| # | Variable |
| Subcategory: Culture (n = 2) |  |
| 1 | Besides English, do you speak or understand another language or dialect? (Youth) |
| 2 | How well do you speak English? (Youth) |
| Subcategory: Family Environment (n = 9) |  |
| 1 | Family members sometimes hit each other. (Youth) |
| 2 | Family members hardly ever lose their tempers. (Youth) |
| 3 | Family members often try to one-up or out-do each other. (Youth) |
| 4 | Family members rarely become openly angry. (Youth) |
| 5 | Family members sometimes get so angry they throw things. (Youth) |
| 6 | Family members often criticize each other. (Youth) |
| 7 | In our family, we believe you don't ever get anywhere by raising your voice. (Youth) |
| 8 | If there's a disagreement in our family, we try hard to smooth things over and keep the peace. (Youth) |
| 9 | We fight a lot in our family. (Youth) |
| Subcategory: Parenting Behavior (n = 16) |  |
| 1 | Is there a second adult who cares for you, that you spend a significant amount of time with, like your other parent, step-paren |
| 2 | (Caregiver 2) Believes in showing their love for me. |
| 3 | (Caregiver 1) Believes in showing their love for me. |
| 4 | (Caregiver 1) Smiles at me very often. |
| 5 | (Caregiver 2) Is able to make me feel better when I am upset. |
| 6 | (Caregiver 2) Makes me feel better after talking over my worries with them. |
| 7 | (Caregiver 1) Makes me feel better after talking over worries with them. |
| 8 | (Caregiver 1) Is able to make me feel better when I am upset. |
| 9 | (Caregiver 1) Is easy to talk to. |
| 10 | (Caregiver 2) Is easy to talk to. |
| 11 | (Caregiver 2) Smiles at me very often. |
| 12 | How often do you talk to your parent or guardian about your plans for the coming day, such as your plans about what will happen |
| 13 | In an average week, how many times do you and your parents/guardians, eat dinner together? |
| 14 | How often do your parents/guardians know where you are? |
| 15 | How often do your parents know who you are with when you are not at school and away from home? |

| Domain: Friends, Family & Community ($n = 48$) | |
| --- | --- |
| # | Variable |
| 16 | If you are at home when your parents or guardians are not, how often do you know how to get in touch with them? |
| Subcategory: Peers ($n = 6$) | |
| 1 | How many of your friends: Have skipped school |
| 2 | How many of your friends: Have shoplifted occasionally |
| 3 | How many of your friends: Are excellent students (GPA 3.5 (B+) or higher) |
| 4 | How many of your friends: Are athletes |
| 5 | How many of your friends: Have been suspended from school |
| 6 | How many of your friends: Attend spiritual or religious services once a month or more often |
| Subcategory: Temperament / Personality ($n = 3$) | |
| 1 | I am helpful if someone is hurt, upset, or feeling sick. |
| 2 | I try to be nice to other people. I care about their feelings. |
| 3 | I often offer to help others (parents, teachers, children). |
| Subcategory: Work and School ($n = 12$) | |
| 1 | In general, I like school a lot. |
| 2 | I get along with my teachers. |
| 3 | Getting good grades is not so important to me. |
| 4 | The school lets my parents know when I have done something well. |
| 5 | There are lots of chances for students in my school to get involved in sports, clubs, or other school activities outside of cla |
| 6 | Usually, school bores me. |
| 7 | I feel I'm just as smart as other kids my age. |
| 8 | I like school because I do well in class. |
| 9 | In my school, students have lots of chances to help decide things like class activities and rules. |
| 10 | My teacher(s) notices when I am doing a good job and lets me know about it. |
| 11 | I feel safe at my school. |
| 12 | There are lots of chances to be part of class discussions or activities. |

Physical Health
| Domain: Physical Health ($n = 8$) | |
| --- | --- |
| # | Variable |
| Subcategory: Anthropometrics and vitals ($n = 4$) | |
| 1 | Waist circumference |
| 2 | Participant wearing a cast or prosthesis |
| 3 | Height |
| 4 | Weight |
| Subcategory: Medical / pain variables ($n = 1$) | |
| 1 | Over the past month, have you had any aches or pains in your body? |
| Subcategory: Physical activity ($n = 3$) | |
| 1 | During the past 7 days, on how many days were you physically active for a total of at least 60 minutes per day? |
| 2 | In an average week when you are in school, on how many days do you go to physical education class? |
| 3 | On how many of the past 7 days did you do exercises to strengthen or tone your muscles, such as push-ups, sit-ups, or weight li |

Mental Health
| Domain: Mental Health (*n* = 253) |  |
| --- | --- |
| # | Variable |
| Subcategory: Anxiety (*n* = 38) |  |
| 1 | Diagnosis: Generalized anxiety disorder (F41.1) - Present (Youth) |
| 2 | Diagnosis: Other specified anxiety disorder, generalized anxiety disorder (impairment does not meet minimum duration) (F41.8) - |
| 3 | Diagnosis: Other specified anxiety disorder, generalized anxiety disorder (impairment does not meet minimum duration) (F41.8) - 1 |
| 4 | Diagnosis: Generalized anxiety disorder (F41.1) - Past (Youth) |
| 5 | Symptom: Worry associated with defined symptom(s) - Past (Youth) |
| 6 | Symptom: Excessive worries across breadth of domains - Past (Youth) |
| 7 | Symptom: Worry associated with defined symptom(s) - Present (Youth) |
| 8 | Symptom: Worrying, duration lasting at least 6 months - Past (Youth) |
| 9 | Symptom: Excessive worries across breadth of domains - Present (Youth) |
| 10 | Symptom: Impairment in functioning due to worries - Present (Youth) |
| 11 | Symptom: Life event or stressor generalized - Present (Youth) |
| 12 | Symptom: Excessive worries more days than not - Past (Youth) |
| 13 | Symptom: Clinically significant distress due to worries - Past (Youth) |
| 14 | Symptom: Impairment in functioning due to worries - Past (Youth) |
| 15 | Symptom: Clinically significant distress due to worries - Present (Youth) |
| 16 | Symptom: Difficulty controlling worries - Present (Youth) |
| 17 | Symptom: Worrying, duration lasting at least 6 months - Present (Youth) |
| 18 | Symptom: Difficulty controlling worries - Past (Youth) |
| 19 | Symptom: Excessive worries more days than not - Present (Youth) |
| 20 | Diagnosis: Social anxiety disorder (F40.10) - Present (Youth) |
| 21 | Diagnosis: Other specified anxiety disorder, social anxiety disorder (impairment does not meet minimum duration) (F41.8) - Pres |
| 22 | Diagnosis: Social anxiety disorder (F40.10) - Past (Youth) |
| 23 | Symptom: Clinically significant distress due to social anxiety - Past (Youth) |
| 24 | Symptom: Social fear is excessive given threat or sociocultural context - Present (Youth) |
| 25 | Symptom: Fear of social situations - Present (Youth) |
| 26 | Symptom: Social situations invariably provoke anxiety - Present (Youth) |
| 27 | Symptom: Social anxiety, duration lasting at least 6 months - Past (Youth) |
| # | Variable |
| 28 | Diagnosis: Other specified anxiety disorder, social anxiety disorder (impairment does not meet minimum duration) (F41.8) - Past |
| 29 | Symptom: Social situations avoided or endured with distress - Present (Youth) |
| 30 | Symptom: Clinically significant distress due to social anxiety - Present (Youth) |
| 31 | Symptom: Social anxiety, duration lasting at least 6 months - Present (Youth) |
| 32 | Symptom: Social situations invariably provoke anxiety - Past (Youth) |
| 33 | Symptom: Social fear is excessive given threat or sociocultural context - Past (Youth) |
| 34 | Symptom: Impairment in functioning due to social anxiety - Past (Youth) |
| 35 | Symptom: Life event or stressor social - Present (Youth) |
| 36 | Symptom: Fear of social situations - Past (Youth) |
| 37 | Symptom: Impairment in functioning due to social anxiety - Present (Youth) |
| 38 | Symptom: Social situations avoided or endured with distress - Past (Youth) |
| Subcategory: Emotion / Positive Effect (*n* = 9) |  |
| 1 | I felt attentive (that is, alert or able to pay attention). |
| 2 | I felt calm. |
| 3 | I felt able to concentrate. |
| 4 | I felt confident. |
| 5 | I felt delighted. |
| 6 | I felt energetic. |
| 7 | I felt interested. |
| 8 | I felt enthusiastic |
| 9 | I felt at ease |
| Subcategory: Harm/ Suicidality (*n* = 47) |  |
| 1 | Symptom: Suicidal attempt - Present (Youth) |
| 2 | Symptom: Suicidal ideation, specific plan - Present (Youth) |
| 3 | Symptom: Suicidal ideation thought of method - Past (Youth) |
| 4 | Diagnosis: No past suicidal ideation or behavior - Past (Youth) |
| 5 | Diagnosis: Suicidal ideation, active intent - Past (Youth) |
| 6 | Symptom: Aborted or interrupted suicide attempts - Past (Youth) |
| 7 | Diagnosis: Interrupted attempt - Present (Youth) |
| 8 | Diagnosis: Self injurious behavior, without suicidal intent - Past (Youth) |
| 9 | Diagnosis: Preparatory actions toward imminent suicidal behavior - Present (Youth) |
| # | Variable |
| 10 | Symptom: Suicidal ideation - Past (Youth) |
| 11 | Symptom: Aborted or interrupted suicide attempts - Present (Youth) |
| 12 | Diagnosis: Suicidal ideation, active intent - Present (Youth) |
| 13 | Diagnosis: Suicide attempt - Past (Youth) |
| 14 | Diagnosis: Aborted attempt - Present (Youth) |
| 15 | Diagnosis: Interrupted attempt - Past (Youth) |
| 16 | Diagnosis: Suicidal ideation, active non-specific - Past (Youth) |
| 17 | Symptom: Suicidal ideation thought of method - Present (Youth) |
| 18 | Diagnosis: Suicidal ideation, passive - Present (Youth) |
| 19 | Diagnosis: Self injurious behavior, without suicidal intent - Present (Youth) |
| 20 | Diagnosis: Suicidal ideation, active method - Present (Youth) |
| 21 | Diagnosis: Suicidal ideation, active plan - Present (Youth) |
| 22 | Diagnosis: Suicidal ideation, active method - Past (Youth) |
| 23 | Symptom: Suicidal attempt - Past (Youth) |
| 24 | Symptom: Suicide attempt, thought could die - Past (Youth) |
| 25 | Symptom: Suicide attempt, thought could die - Present (Youth) |
| 26 | Symptom: Number of suicide attempts - Past (Youth) |
| 27 | Symptom: Suicidal ideation, intent to act - Present (Youth) |
| 28 | Diagnosis: Suicide attempt - Present (Youth) |
| 29 | Symptom: Wishes/better off dead - Past (Youth) |
| 30 | Symptom: Self-injury, thought could die from behavior - Past (Youth) |
| 31 | Symptom: Suicidal ideation, specific plan - Past (Youth) |
| 32 | Diagnosis: Suicidal ideation, active non-specific - Present (Youth) |
| 33 | Symptom: Suicidal ideation - Present (Youth) |
| 34 | Symptom: Suicide attempt, method - Present (Youth) |
| 35 | Symptom: Wishes/better off dead - Present (Youth) |
| 36 | Symptom: Self injurious behavior - Past (Youth) |
| 37 | Symptom: Self-injury, thought could die from behavior - Present (Youth) |
| 38 | Symptom: Self injurious behavior - Present (Youth) |
| 39 | Diagnosis: No suicidal ideation or behavior - Present (Youth) |
| 40 | Symptom: Suicidal behavior, made preparations - Past (Youth) |
| # | Variable |
| 41 | Diagnosis: Suicidal ideation, passive - Past (Youth) |
| 42 | Symptom: Suicidal behavior, made preparations - Present (Youth) |
| 43 | Symptom: Suicidal ideation, intent to act - Past (Youth) |
| 44 | Diagnosis: Suicidal ideation, active plan - Past (Youth) |
| 45 | Diagnosis: Preparatory actions toward imminent suicidal behavior - Past (Youth) |
| 46 | Diagnosis: Aborted attempt - Past (Youth) |
| 47 | Symptom: Suicide attempt, method - Past (Youth) |
| Subcategory: Mood / Depressive Disorder (*n* = 49) |  |
| 1 | Symptom: Depressed mood - Present (Youth) |
| 2 | Symptom: Guilt - Present (Youth) |
| 3 | Symptom: Decreased appetite - Past (Youth) |
| 4 | Symptom: Anhedonia - Present (Youth) |
| 5 | Symptom: Concentration disturbance - Past (Youth) |
| 6 | Symptom: Insomnia when depressed - Past (Youth) |
| 7 | Symptom: Weight loss - Past (Youth) |
| 8 | Symptom: Decreased self-esteem - Present (Youth) |
| 9 | Symptom: Indecision - Past (Youth) |
| 10 | Symptom: Anhedonia - Past (Youth) |
| 11 | Diagnosis: Persistent depressive disorder (Dysthymia) (F34.1) - Partial remission (Youth) |
| 12 | Diagnosis: Other specified depressive disorder, persistent depressive disorder (impairment does not meet full criteria) (F32.8) |
| 13 | Diagnosis: Unspecified depressive disorder (F32.9) - Past (Youth) |
| 14 | Symptom: Depressive disorders, no 2-month symptom-free interval - Present (Youth) |
| 15 | Symptom: Increased appetite - Present (Youth) |
| 16 | Symptom: Irritability - Past (Youth) |
| 17 | Symptom: Decreased appetite - Present (Youth) |
| 18 | Symptom: Impairment in functioning due to depression - Past (Youth) |
| 19 | Symptom: Psychomotor agitation in depressive disorder - Present (Youth) |
| 20 | Symptom: Decreased self-esteem - Past (Youth) |
| 21 | Symptom: Depressed mood - Past (Youth) |
| 22 | Diagnosis: Major depressive disorder (F32.4) - Partial remission (Youth) |
| 23 | Symptom: Presence of two or more depressive symptoms - Present (Youth) |
| # | Variable |
| 24 | Symptom: Weight gain - Present (Youth) |
| 25 | Symptom: Hypersomnia - Past (Youth) |
| 26 | Diagnosis: Persistent depressive disorder (Dysthymia) (F34.1) - Past (Youth) |
| 27 | Symptom: Hypersomnia - Present (Youth) |
| 28 | Symptom: Hopeless - Past (Youth) |
| 29 | Symptom: Presence of two or more depressive symptoms - Past (Youth) |
| 30 | Diagnosis: Major depressive disorder (F32.9) - Past (Youth) |
| 31 | Symptom: Psychomotor retardation - Past (Youth) |
| 32 | Symptom: Indecision - Present (Youth) |
| 33 | Diagnosis: Major depressive disorder - Present (Youth) |
| 34 | Symptom: Psychomotor retardation - Present (Youth) |
| 35 | Symptom: Hopeless - Present (Youth) |
| 36 | Symptom: Depressive disorders, no 2-month symptom-free interval - Past (Youth) |
| 37 | Diagnosis: Unspecified depressive disorder (F32.9) - Present (Youth) |
| 38 | Symptom: Fatigue - Present (Youth) |
| 39 | Diagnosis: Persistent depressive disorder (Dysthymia) (F34.1) - Present (Youth) |
| 40 | Symptom: Irritability - Present (Youth) |
| 41 | Symptom: Life event or stressor - Present (Youth) |
| 42 | Symptom: Weight loss - Present (Youth) |
| 43 | Symptom: Guilt - Past (Youth) |
| 44 | Symptom: Fatigue - Past (Youth) |
| 45 | Symptom: Increased appetite - Past (Youth) |
| 46 | Symptom: Impairment in functioning due to depression - Present (Youth) |
| 47 | Symptom: Weight gain - Past (Youth) |
| 48 | Symptom: Concentration disturbance - Present (Youth) |
| 49 | Symptom: Psychomotor agitation in depressive disorder - Past (Youth) |
| Subcategory: Peer Behavior (*n* = 19) |  |
| 1 | Youth's total number of close friends |
| 2 | A kid tried to damage my social reputation by spreading rumors about me |
| 3 | I left another kid out of what I was doing. |
| 4 | I said mean things about a kid so that people would think they were a loser. |
| # | Variable |
| 5 | I threatened to hurt or beat up another kid. |
| 6 | A kid hit, kicked, or pushed me in a mean way. |
| 7 | I chased a kid like I was really trying to hurt them. |
| 8 | A kid did not invite me to a party or social event though they knew that I wanted to go. |
| 9 | Another kid said mean things about me so that people would think I was a loser. |
| 10 | I gossiped about another kid so others would not like them. |
| 11 | I hit, kicked, or pushed another kid in a mean way. |
| 12 | A kid left me out of what they were doing. |
| 13 | Some kids left me out of an activity or conversation I really wanted to be included in. |
| 14 | A kid chased me like they were really trying to hurt me. |
| 15 | Another kid gossiped about me so others would not like me. |
| 16 | I did not invite a kid to a party or other social event even though I knew the kid wanted to go. |
| 17 | A kid threatened to hurt or beat me up. |
| 18 | I left another kid out of an activity or conversation that they really wanted to be included in. |
| 19 | I tried to damage another kid's social reputation by spreading rumors about them. |
| Subcategory: Psychotic Behavior (*n* = 21) |  |
| 1 | Did you ever feel very certain that you have very special abilities or magical talents that other people do not have? |
| 2 | Did places that you know well, such as your bedroom, or other rooms in your home, your classroom or school yard, suddenly seem |
| 3 | Did you feel like you had special, unusual powers like you could make things happen by magic, or that you could magically know |
| 4 | Did things you looked at seem different than they usually do; like did they seem shinier or darker, larger or smaller or change |
| 5 | Did you feel that parts of your body had suddenly changed or worked differently than before; like your legs had suddenly turned |
| 6 | Did you suddenly find it hard to figure out how to say something quickly and easily so that other people would understand what |
| 7 | Did you suddenly start to notice that people sometimes had a hard time understanding what you were saying, even though they use |
| 8 | Did you feel that the world is not real, you are not real, or that you are dead? |
| 9 | Although you could not see anything or anyone, did you suddenly start to feel that an invisible energy, creature, or some perso |
| 10 | Did you start to worry at times that your mind was trying to trick you or was not working right? |
| # | Variable |
| 11 | Did you feel confused because something you experienced didn't seem real or it seemed imaginary to you? |
| 12 | Did you feel that other people might want something bad to happen to you or that you could not trust other people? |
| 13 | Did you suddenly feel that you could not trust other people because they seemed to be watching you or talking about you in an u |
| 14 | Did your skin or just beneath your skin suddenly start feeling strange, like bugs crawling? |
| 15 | Did you honestly believe in things that other people would say are unusual or weird? |
| 16 | Did you feel that sometimes your thoughts were so strong you could almost hear them, as if another person, not you, spoke them? |
| 17 | Did you feel that someone else, who is not you, has taken control over the private, personal, thoughts or ideas inside your hea |
| 18 | Did you hear strange sounds that you never noticed before like banging, clicking, hissing, clapping, or ringing in your ears? |
| 19 | Did you suddenly start to be able to see things that other people could not see or they did not seem to see? |
| 20 | Did you lose concentration because you noticed sounds in the distance that you usually don't hear? |
| 21 | Did you suddenly start to see unusual things that you never saw before like flashes, flames, blinding light, or shapes floating |
| Subcategory: Sleep Problems (*n* = 4) |  |
| 1 | Symptom: Insomnia - Present (Youth) |
| 2 | Diagnosis: Sleep problems - Present (Youth) |
| 3 | Symptom: Insomnia - Past (Youth) |
| 4 | Diagnosis: Sleep problems - Past (Youth) |
| Subcategory: Stress Levels (*n* = 25) |  |
| 1 | In your life time - Someone in family died? (Youth) |
| 2 | In your life time - Got new step-parent? (Youth) |
| 3 | In your life time - Sibling left home? (Youth) |
| 4 | In your life time - Was a victim of crime/violence/assault? (Youth) |
| 5 | In your life time - Family member had mental/emotional problem? (Youth) |
| 6 | In your life time - Negative change in parent's financial situation? (Youth) |
| 7 | In your life time - One of the parents/caregivers went to jail? (Youth) |
| 8 | In your life time - Parents separated or divorced? (Youth) |
| 9 | In your life time - Someone in the family was arrested? (Youth) |
| # | Variable |
| 10 | In your life time - Parental figure lost job? (Youth) |
| 11 | In your life time - Family member was seriously injured? (Youth) |
| 12 | In your life time - You got seriously injured? (Youth) |
| 13 | In your life time - You got seriously sick? (Youth) |
| 14 | In your life time - Family moved? (Youth) |
| 15 | In your life time - Family member had drug and/or alcohol problem? (Youth) |
| 16 | In your life time - Attended a new school? (Youth) |
| 17 | In your life time - Got new sibling? (Youth) |
| 18 | In your life time - Close friend was seriously sick/injured? (Youth) |
| 19 | In your life time - Saw crime or accident? (Youth) |
| 20 | In your life time - Parents/caregiver got into trouble with the law? (Youth) |
| 21 | In your life time - Parent/caregiver got a new job? (Youth) |
| 22 | In your life time - One parent was away from home more often? (Youth) |
| 23 | In your life time - Lost a close friend? (Youth) |
| 24 | In your life time - Parents argued more than previously? (Youth) |
| 25 | In your life time - Close friend died? (Youth) |
| Subcategory: Temperament / Personality (*n* = 41) |  |
| 1 | I am very fearful compared to my friends. |
| 2 | I often do things for no other reason than they might be fun. |
| 3 | When I see an opportunity to get something I want, I go for it right away. |
| 4 | I feel excited and full of energy when I get something that I want. |
| 5 | It would excite me to win a contest. |
| 6 | I do not become fearful or nervous, even when something bad happens to me. (Parent) |
| 7 | I get thrilled when good things happen to me. |
| 8 | When I am doing well at something, I like to keep doing this. |
| 9 | When I want something, I usually go all the way to get it. |
| 10 | Nobody can stop me when I want something. |
| 11 | I get really excited when I see an opportunity to get something I like. |
| 12 | I am always willing to try something new, when I think it will be fun. |
| 13 | I do everything to get the things that I want. |
| 14 | I often do things on the spur of the moment. |
| # | Variable |
| 15 | I am hurt when people scold me or tell me that I do something wrong. |
| 16 | I feel pretty upset when I think that someone is angry with me. |
| 17 | I feel worried when I have done poorly at something. |
| 18 | I worry about making mistakes. |
| 19 | I do not become fearful or nervous, even when something bad happens to me. (Youth; Version 1) |
| 20 | I crave excitement and new sensations. |
| 21 | I usually get very tense when I think something unpleasant is going to happen. |
| 22 | I am very careful. |
| 23 | I tend to lose control when I am in a great mood. |
| 24 | I would like to learn to fly an airplane. |
| 25 | When I get really happy about something, I tend to do things that can lead to trouble. |
| 26 | I like to stop and think about things before I do it. (Tendency to think carefully before acting) |
| 27 | I would like to ski very fast down a high mountain slope. |
| 28 | I almost always finish projects that I start. |
| 29 | I enjoy taking risks. |
| 30 | When I am in a great mood, I tend to do things that can cause me problems. |
| 31 | I am a person who always gets the job done. |
| 32 | I tend to act without thinking when I am very, very happy. |
| 33 | I like new, thrilling things, even if they are a little scary. |
| 34 | When I feel bad, I often do things I later regret in order to make myself feel better now. |
| 35 | Sometimes when I feel bad, I keep doing something even though it is making me feel worse. |
| 36 | I tend to get things done on time. |
| 37 | When I feel rejected, I often say things that I later regret. |
| 38 | I tend to stop and think before doing things. |
| 39 | I try to take a careful approach to things. |
| 40 | I finish what I start. |
| 41 | When I am upset I often act without thinking. |

